# Potentially causal associations between placental DNA methylation and schizophrenia and other neuropsychiatric disorders

**DOI:** 10.1101/2023.03.07.23286905

**Authors:** Ariadna Cilleros-Portet, Corina Lesseur, Sergi Marí, Marta Cosin-Tomas, Manuel Lozano, Amaia Irizar, Amber Burt, Iraia García-Santisteban, Diego Garrido Martín, Geòrgia Escaramís, Alba Hernangomez-Laderas, Raquel Soler-Blasco, Charles E. Breeze, Bárbara P. Gonzalez-Garcia, Loreto Santa-Marina, Jia Chen, Sabrina Llop, Mariana F. Fernández, Martine Vrijhed, Jesús Ibarluzea, Mònica Guxens, Carmen Marsit, Mariona Bustamante, Jose Ramon Bilbao, Nora Fernandez-Jimenez

## Abstract

Increasing evidence supports the role of placenta in neurodevelopment and potentially, in the later onset of neuropsychiatric disorders. Recently, methylation quantitative trait loci (mQTL) and interaction QTL (iQTL) maps have proven useful to understand SNP-genome wide association study (GWAS) relationships, otherwise missed by conventional expression QTLs. In this context, we propose that part of the genetic predisposition to complex neuropsychiatric disorders acts through placental DNA methylation (DNAm). We constructed the first public placental *cis*-mQTL database including nearly eight million mQTLs calculated in 368 fetal placenta DNA samples from the INMA project, ran cell type- and gestational age-imQTL models and combined those data with the summary statistics of the largest GWAS on 10 neuropsychiatric disorders using Summary-based Mendelian Randomization (SMR) and colocalization. Finally, we evaluated the influence of the DNAm sites identified on placental gene expression in the RICHS cohort. We found that placental *cis*-mQTLs are highly enriched in placenta-specific active chromatin regions, and useful to map the etiology of neuropsychiatric disorders at prenatal stages. Specifically, part of the genetic burden for schizophrenia, bipolar disorder and major depressive disorder confers risk through placental DNAm. The potential causality of several of the observed associations is reinforced by secondary association signals identified in conditional analyses, regional pleiotropic methylation signals associated to the same disorder, and cell type- imQTLs, additionally associated to the expression levels of relevant immune genes in placenta. In conclusion, the genetic risk of several neuropsychiatric disorders could operate, at least in part, through DNAm and associated gene expression in placenta.

## Introduction

The impact of the intrauterine environment on development and health, both fetal and long-term, has been known for many years^1^. In this line, the developmental origins of health and disease (DOHaD) hypothesis postulates that perinatal and early life environments can impact fetal and later-life health^2^. Prenatal stress affects the quality of the intrauterine environment, and is highly related to cardiovascular and metabolic, as well as behavioral and neurodevelopmental disorders^3^. Thus, the placenta is an ephemeral fetal organ that is uniquely situated to evaluate prenatal exposures in the context of DOHaD, because it manages the transport of nutrients, oxygen, waste, and endocrine signals between mother and fetus^4^. Additionally, the placenta constitutes the interface between mother and child during pregnancy and is the maximum regulator of the prenatal milieu, having a key role in the growth and the neurodevelopment of the fetus^5^. Hence, the placenta has been described as the third brain that links the fetal brain with the mature maternal brain, thus becoming the cornerstone to understand the prenatal environmental effects on neurodevelopment^6^, and potentially, on the appearance of neurodevelopmental and neuropsychiatric disorders later in life.

Particularly in the case of schizophrenia (SCZ), many susceptibility genes have been identified, demonstrating a remarkable genetic basis, but a considerable amount of research suggest that environmental factors may also play a role^7^. The neurodevelopmental hypothesis of SCZ was first proposed by Daniel Weinberger in 1987 and has been reinforced in the last decades, with increasing evidence of neurodevelopmental abnormalities contributing to the pathophysiology of the disease^8,9,10^. The central argument of this hypothesis states that abnormal fetal neurodevelopment creates a vulnerability to develop SCZ later in life. In fact, experimental evidence states that prenatal insults such as maternal immune activation (MIA) are associated with SCZ in offspring^11^. MIA occurs when inflammatory markers rise above the normal range in pregnancy as a result of maternal inflammation and can be caused by psychosocial stress, infection, or other factors^12^.

Besides, there are many studies proposing that the intrauterine environment alters the placental function through epigenetic mechanisms, such as DNA methylation (DNAm)^13^. It is very important to note that, while DNAm is bimodally distributed in most tissues and cell types, it follows a trimodal distribution in the very specific case of placenta, due to its high content of both partially methylated domains and CpG positions with intermediate methylation levels^14^. Although a very recent study has pointed out that DNAm of healthy donors could be highly cell type-specific, with modest impact of either genetics or other factors^15^, it is worth to mention that DNAm has been considered as a bridge between the environment and the genome that at the same time, is under the control of both environmental and genetic factors. The peculiarity of placenta as an ephemeral organ that connects two organisms, together with its very particular methylome that is remarkably enriched in CpG sites with intermediate methylation levels^14^, leads us to speculate that the genome-environment interaction and its impact on DNAm is very likely unique in placenta and deserves further investigation.

In the past several years, different works have demonstrated that placental DNAm is sensitive to environmental factors surrounding the gestation. In 2021 the Pregnancy and Childhood Epigenetics (PACE) consortium conducted a meta-analysis on 1,700 placental samples and found a placenta-specific DNAm signature of maternal smoking during pregnancy, with differentially methylated CpG sites located in active regions of the placental epigenome, and close to genes involved in the regulation of inflammatory activity, growth factor signaling, and response to environmental stressors^16^. One year later, the same consortium reported in the largest placental DNAm study conducted to date that maternal pre-pregnancy body mass index also impacts the placental methylome, specifically at CpG sites located close to obesity-related genes and altogether, enriched in oxidative-stress and cancer pathways^17^.

Noting the impact of the genetic background on placental DNAm, several studies have reported a number of methylation quantitative trait loci (mQTL) in placenta and have used them in different downstream applications^18,19,20,21^. Among others, Tekola-Ayele and collaborators showed candidate functional pathways that underpin the genetic regulation of birth weight via placental epigenetic and transcriptomic mechanisms^20^. However, the catalogue of mQTLs they provided was limited to birth weight-related genomic loci. More recently, Casazza reported nearly 50,000 placental *cis*-mQTLs and 2,489 sex-specific placental *cis*-mQTLs^21^. They found out that placental mQTLs were enriched in genome-wide association study (GWAS) loci for both growth- and immune-related traits, and that male- and female-specific mQTLs were more abundant than non-sex-specific ones. Another remarkable discovery is that they found a modest enrichment of placental mQTLs in proportion to the calculated SNP heritability in the case of neuropsychiatric disorders compared to immune- and growth-related traits. However, they did not rule out the possibility that a part of the genetic susceptibility of neuropsychiatric disorders is conferred through modification of the placenta, and if this was the case, it would be relevant not only in terms of etiopathology, but also from a clinical viewpoint, due to the importance of localizing therapeutic targets in the right tissues, contexts and developmental stages.

In cross-tissue analyses, mQTLs have been highlighted as powerful instruments revealing molecular links to traits otherwise missed by expression quantitative trait loci (eQTL). In fact, recent work by Oliva et al. reported that trait-associated variants are more likely to result in detectable changes in DNAm rather than gene expression, highlighting the relevance of multi-tissue mQTL maps^22^. On the other hand, in 2021, cell type-interacting eQTLs were defined as proxies of cell type-specific eQTLs and were found not to be covered by standard eQTLs, while they appeared to be highly valuable for gaining a mechanistic understanding of complex trait associations^23^. Taken together, placenta-specific mQTLs as well as placenta cell type interacting mQTLs (imQTLs) can be useful tools in the search for the etiology of genetic associations. In this sense, it is of outmost importance to make this type of resources public and available to the scientific community.

Taking into consideration i) the increasing evidence supporting the role of placenta in neurodevelopment and potentially, in the onset of neuropsychiatric traits and disorders, ii) the peculiarity of the placental methylome, and iii) the potential of multi-tissue mQTL and imQTL maps to clarify the etiology of complex traits, we proposed that part of the genetic predisposition to complex neuropsychiatric disorders acts through the placental methylome. Thus, we constructed the first publicly available placental *cis*-mQTL database including nearly 8 million nominal mQTLs calculated in 368 fetal placenta samples, investigated cell type- and gestational age (GA)-imQTLs and integrated all these data with summary statistics of the largest GWAS on 10 neuropsychiatric disorders, using Summary-based Mendelian Randomization (SMR) and colocalization approaches. Finally, we evaluated the functional role of the identified DNAm sites on placental gene expression. We found that placental *cis*-mQTLs are enriched in placental active genomic regions, and are useful to map the etiology of neuropsychiatric disorders to prenatal stages. Specifically, part of the SCZ, bipolar disorder (BIP) and major depressive disorder (MDD) genetic risk could act through placental DNAm. The potential causality of several associations observed is reinforced by secondary association signals identified in conditional analyses, regional pleiotropic methylation signals associated to the same disorder, as well as cell type imQTLs involved, that additionally associate to the expression levels of relevant genes in placenta.

## Results

### Placental *cis*-mQTL characterization

The nominal mQTL database was calculated in 368 fetal placenta DNA samples from the Gipuzkoa, Sabadell and València cohorts of the *Infancia y Medio Ambiente* (INMA) project^24^, and contained 7,921,914 *cis*-mQTLs (P_nominal_ < 5×10^−8^) with 110,721 and 1,900,580 unique CpGs and SNPs, respectively. Briefly, placental DNAm was modeled as a function of genotype in 0.5 Mb windows, with fetal sex, 5 genotype principal components (PC) and Planet-estimated cell types^25^ as covariates. Phenotype data of the donor mothers is summarized in Table 1. The vast majority of the SNP-CpG pairs in *cis*-mQTLs were located close to each other with a median distance of 44 kb, indicating that genetically modulated DNAm is typically close to the implicated regulatory variant, as previously described by other authors^26^ (Figure 1A). The distribution of mQTLs across chromosomes was in line with chromosome length, except for chromosome 6, where a higher numer of CpG-SNP pairs was observed (883,548 CpG-SNP pairs with 8,173 and 153,743 unique CpGs and SNPs, respectively) (Figure 1B). This was expected due to the complex linkage disequilibrium (LD) structure of the Human Leucocyte Antigen (HLA) region on this chromosome. Additionally, placental *cis*-mQTLs presented a uniform distribution of effect sizes and directions (Figure 1C).

**Table 1.**
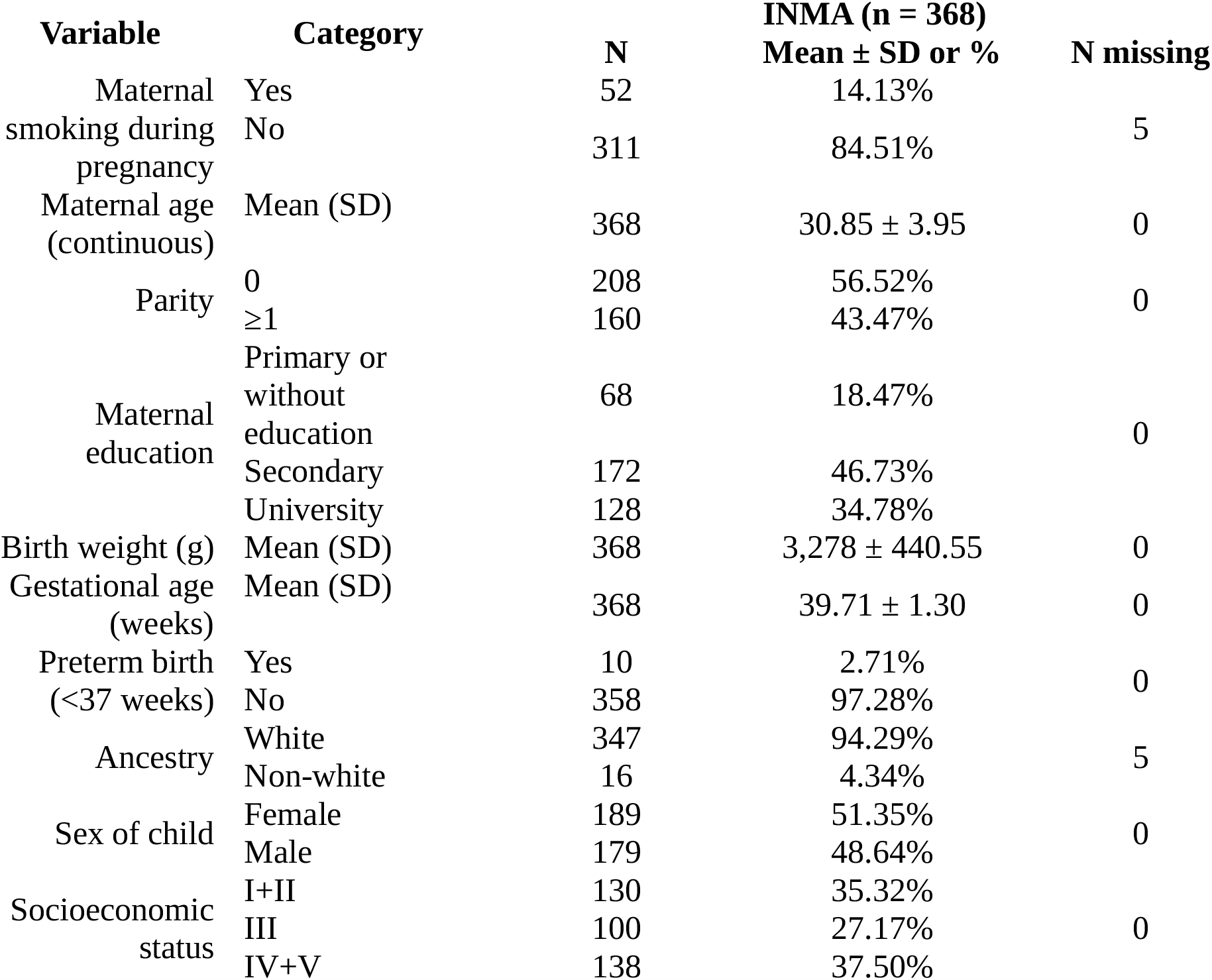
Distribution of maternal smoking during pregnancy, demographic variables, birth outcomes and covariates of the INMA cohort. SD = Standard Deviation.

**Figure 1.**
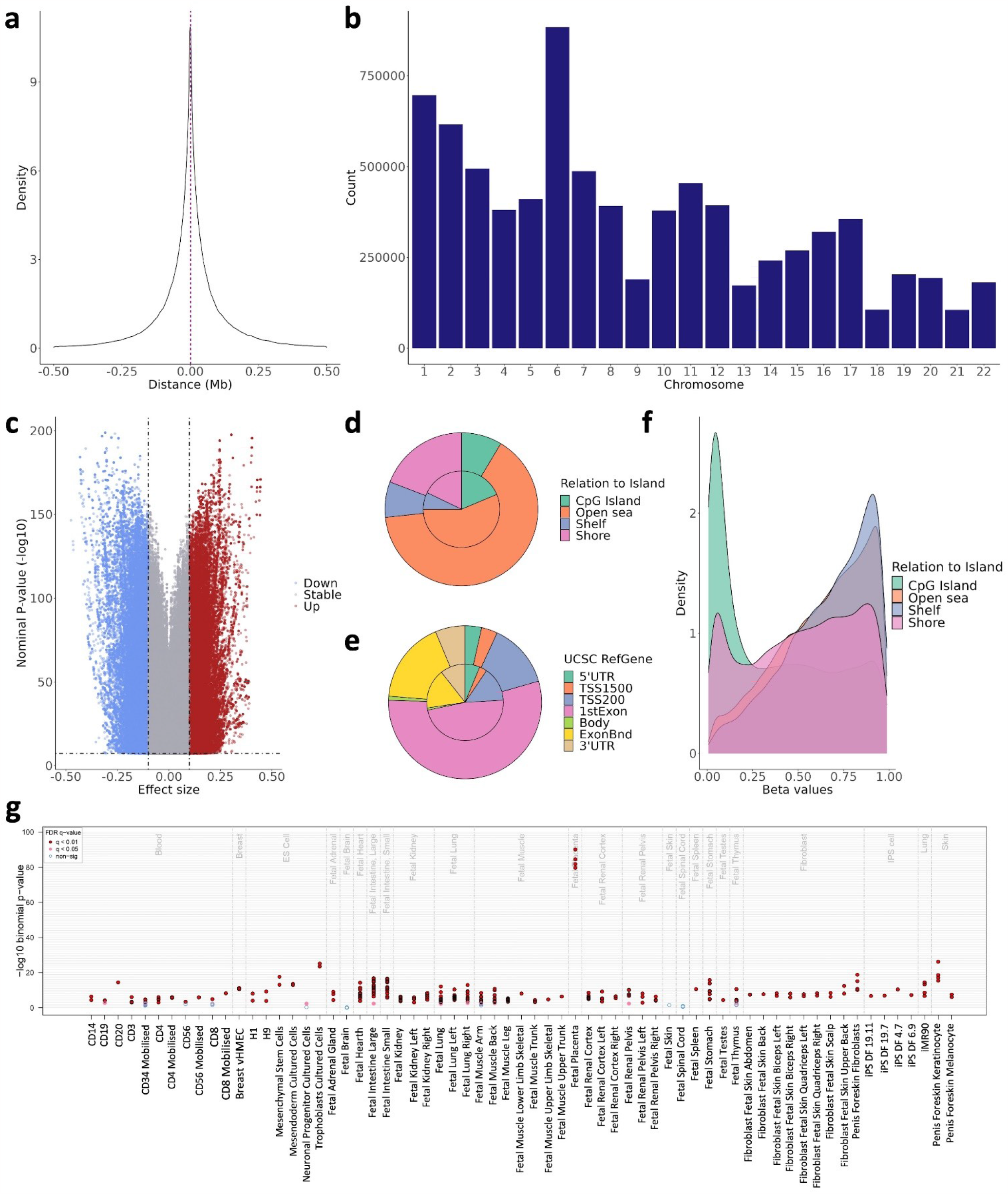
Characterization of the placental *cis*-mQTLs from the nominal database. The distance between the SNP-CpG pair participating in the reported *cis*-mQTLs is displayed as **a** density plot, where the X-axis represents the distance in Mb. The red line represents the median distance of 44 kb. The distribution of the reported *cis*-mQTLs along the chromosomes is shown in the **b** barplot, where the X-axis represents the autosomal chromosomes. The uniform distribution of the effect size from the reported *cis-*mQTLs is pictured in the **c** volcano plot, where the Y-axis illustrates the -log10 nominal p-value, and the X-axis the effect size. The blue and the red dots represent the mQTLs with a negative and positive effect, respectively. The distribution of the EPIC array probes (inner circle) and the nominal mQTL-CpGs (outer circle) considering the Relation To Island and the UCSC RefGene annotation is displayed in the **d** and **e** piecharts, respectively. The methylation beta values, ranging from 0 to 1, of the participating mQTL-CpGs stratified by the Relation To Island annotation is shown in the **f** density plot, where the methylation values are found in the X-axis. The eFORGE enrichment of DNase I hotspots considering the top 10,000 nominal mQTL-CpGs is shown in the **g** plot. The Y-axis represents the -log_10_ binomial p-value of the enrichment, and the X-axis to the tissue. False Discovery Rate (FDR) corrected q-values below 0.01 and 0.05 are represented by red and pink dots, respectively, while blue dots show q-values >0.05.

In general, mQTL-CpGs were depleted from CpG islands and promoters (P < 2.2×10^−16^ and P < 2.2×10^−16^, respectively), and enriched within gene bodies (P = 1.71×10^−8^) and genomic features showing intermediate methylation values such as open sea, and CpG island shelf and shore regions (P < 2.2×10^−16^, P = 3.83×10^−7^ and P < 2.2×10^−16^, respectively) (Figure 1D, E and F, and Supplementary Data 1). Using eFORGE^27,28,29^, we were able to detect an enrichment in fetal placenta-specific DNase I hotspots (Figure 1G) and H3K4me1 broadPeaks (Supplementary Figure 1A and Supplementary Data 2), that mark accessible chromatin and enhancer regions, respectively. Afterwards, we annotated the mQTL-CpGs to their closest genes, performed a gene set enrichment analysis with the Disease Ontology database^30^, and obtained 140 enriched gene sets, including developmental disorder of mental health (Benjamini-Hochberg P = 1.41×10^−10^), psychotic disorder (Benjamini-Hochberg P = 1.41×10^−10^) and SCZ (Benjamini-Hochberg P = 1.41×10^−10^) (Supplementary Data 3 and Supplementary Figure 2).

Apart from the nominal *cis*-mQTLs, permuted and conditional *cis*-mQTLs were also calculated in our 368 placenta DNA samples. Both permuted and conditional mQTL databases showed a median SNP-CpG distance of 7kb, remarkably shorter than in the nominal database. Chromosome 6 was no longer the one showing the highest absolute number of *cis*-mQTLs (Supplementary Figures 3 and 4). These two observations were expected considering that both permuted and conditional mQTL analyses give as a result a reduced number of SNPs per CpG compared to the nominal approach. Additionally, we observed enrichments and depletions similar to those reported for the nominal database (Supplementary Figures 1, 3 and 4, and Supplementary Data 1, 2 and 3). When we compared the *cis*-mQTLs included in the three different databases, the vast majority of the mQTLs were common. The nominal database computes all the possible mQTLs in the SNP-CpG window. Therefore, as expected, it contained a much larger number of mQTLs than the two other databases (Supplementary Figure 5).

The three complete placental *cis*-mQTL databases are publicly available online in the following address: https://smari.shinyapps.io/shi

### Placental cell type- and GA-imQTLs

The most abundant cell type in fetal placenta are syncytiotrophoblasts (STB). During gestation, undifferentiated trophoblasts (TB) change into fully differentiated STB, a continuous, specialized layer of epithelial cells^31^. We estimated the cell-type proportions of our samples by using the reference-based method Planet^25^. As expected, the estimated content of STB was negatively correlated with the estimated proportion of TB in our placental samples (P < 2.2×10^−16^, R^2^ = -0.89). In turn, STB content was positively correlated with gestational age (GA) (P < 4.5×10^−5^, R^2^ = 0.21) (Figure 2A and 2B). We calculated STB-, TB- and GA-imQTLs, as proxies for STB-, TB- and GA-specific *cis*-mQTLs, and obtained 202 positive and 728 negative STB-imQTLs, 237 positive and 594 negative TB-imQTLs, and 412 GA-imQTLs (P_interaction_ < 5×10^−8^) (Supplementary Data 4). The higher amount of STB-imQTLs, revealed a higher statistical power for the most abundant cell type, as previously stated by Kim-Hellmuth et al^23^. Out of the 930 STB-imQTLs and 831 TB-imQTLs, 443 were common to both cell types (Figure 2C). Considering the same allele for both STB and TB, the effect sizes of the shared cell type-imQTLs were negatively correlated (P < 2.2×10^−16^, R^2^ = -0.98) with opposite effect directions as in the example in Figure 2D, E and F. Of note, most of the overlapping imQTLs were positively correlated with the STB content, and negatively correlated with the TB abundance (431/443). In contrast, no overlap was observed with GA-imQTLs (Figure 2C). The effect of GA on the genetic regulation of placental DNAm seemed to be independent of the cell type composition of each sample and therefore, rather a matter of time of gestation regardless of the TB to STB transition.

**Figure 2.**
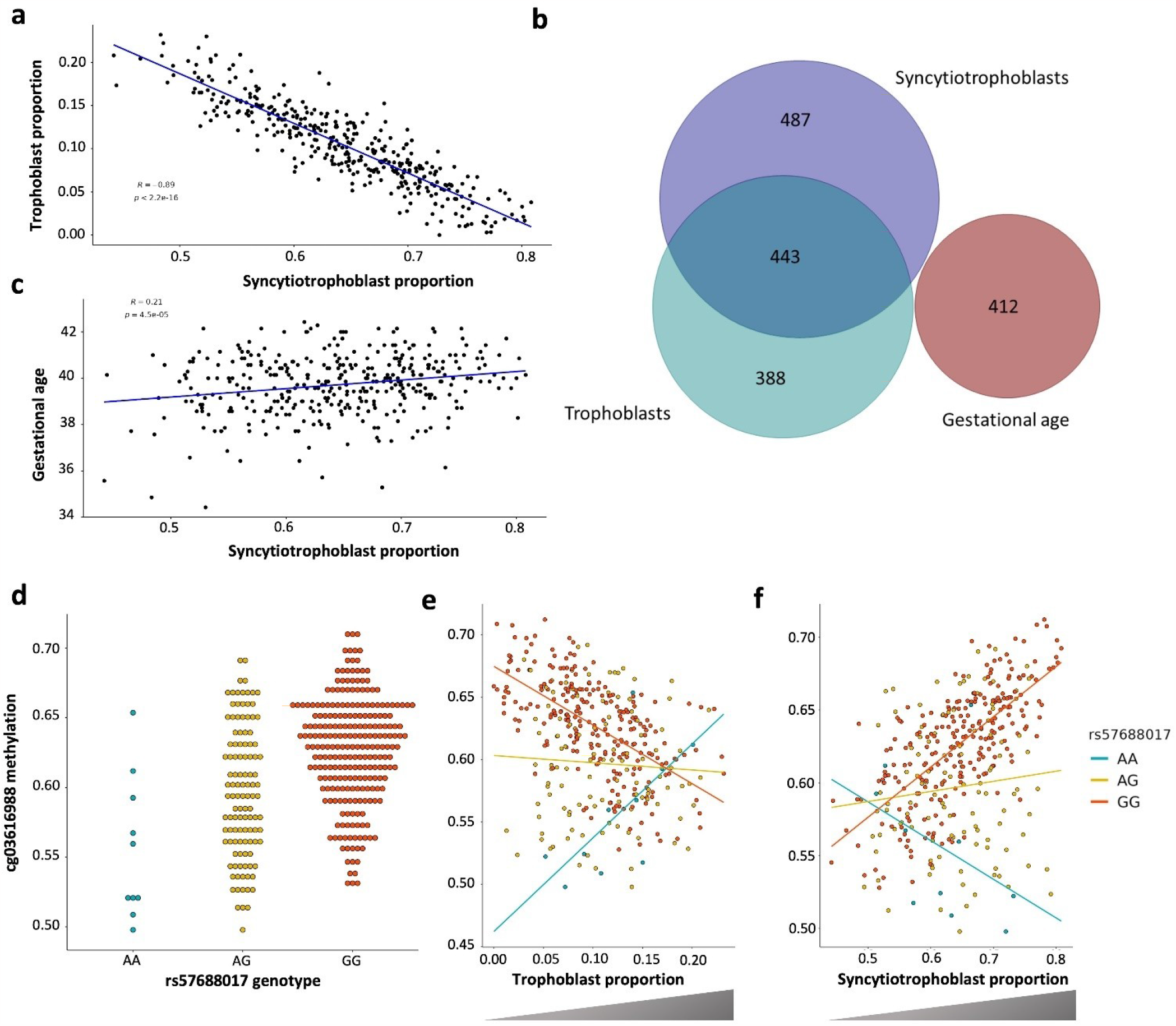
STB-, TB- and GA-imQTLs. The correlation between STB and TB cell types (N=368) is shown in **a**, the X-axis representing the STB proportion in the sample set, and the Y-axis showing the estimated TB proportion. The intersection between STB-, TB- and GA-imQTLs is represented in the **b** Venn diagram. The correlation between STB and GA (N=368) is shown in **c**, the X-axis representing the STB proportion in the sample set, and the Y-axis showing the GA. The standard cg03616988-rs57688017 mQTL, as well as the TB- and STB-imQTLs, are displayed in the **d, e** and **f** dotplots, respectively. In all three, the Y-axes represent the cg03616988 DNAm beta values, ranging from 0 to 1. In **d**, the X-axis displays the genotype of rs57688017, while in **e** and **f**, the X-axes show the TB and STB proportion, respectively.

**Figure 3.**
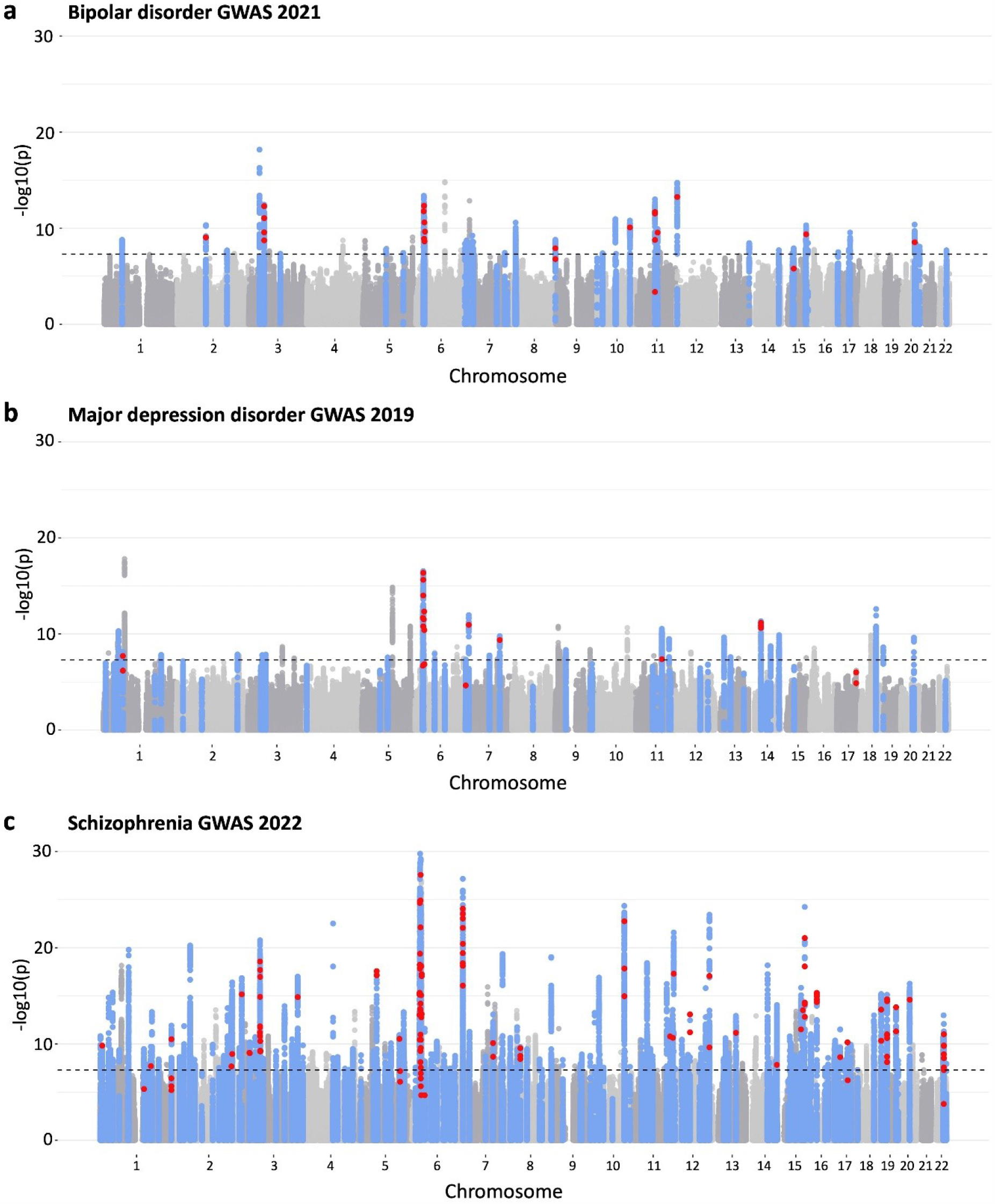
Manhattan plot of BIP, MDD and SCZ GWAS highlighting the SMR and *coloc* results. The original GWAS from BIP, MDD and SCZ were plotted in the **a, b** and **c** Manhattan plots, respectively. In the Y-axes the original -log_10_ p-values are displayed, and in the X-axis the chromosomes. The blue dots represent genomic regions significantly colocalizing with our placental mQTLs, and the red dots are mQTL-SNPs associated with CpGs that have been shown to pleiotropically associate with either BIP, MDD or SCZ in the SMR approach. Therefore, the blue dots represent the colocalization results, and the red dots show the SMR results.

### Multi-omics approaches to unravel the placental origin of neuropsychiatric traits and disorders

We selected the largest GWAS for each of the traits analyzed, namely attention deficit and hyperactivity disorder (ADHD)^32^, aggressive behavior in children (AGR)^33^, autism spectrum disorder (ASD)^34^, BIP^35^, internalizing problems (INT)^36^, MDD^37^, obsessive-compulsive disorder (OCD)^38^, panic disorder (PD)^39^, suicidal attempt (SA)^40^ and SCZ^41^ (Table 2). Most of the studies were carried out in the context of either the Psychiatric Genomics Consortium (PGC)^42^ or the Early Genetics and Lifecourse Epidemiology (EAGLE)^43^ consortium. GWAS sample sizes ranged from 6,183 in the case of SA and 500,199 samples in MDD (Table 2).

**Table 2.**
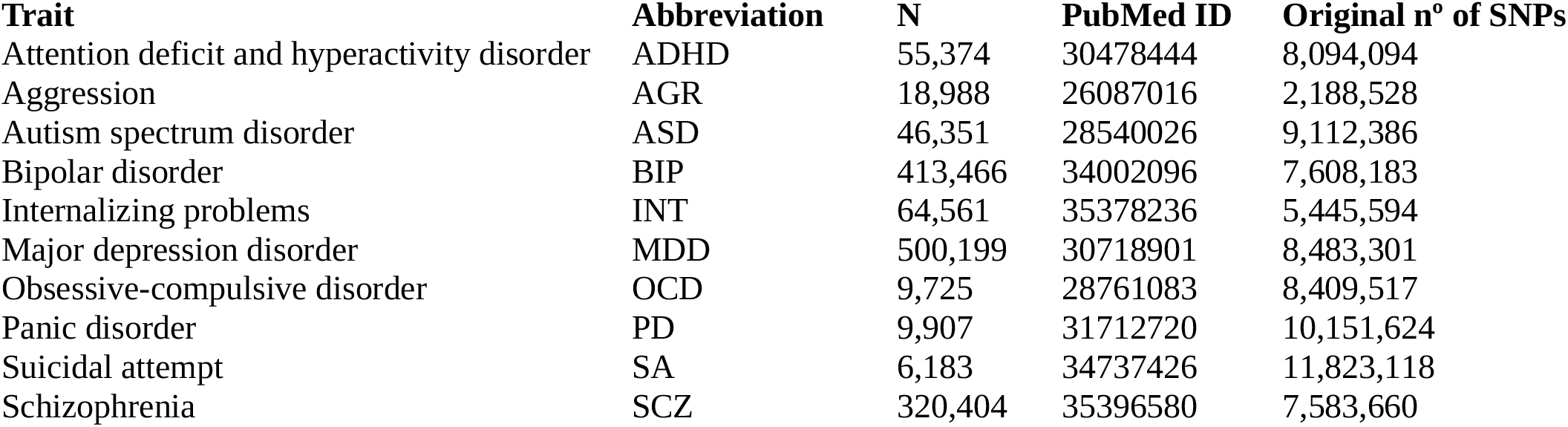
Overview of the neuropsychiatric traits and disorders included in this study.

We searched for placenta DNAm sites pleiotropically associated with neuropsychiatric disorders using the SMR and the heterogeneity in dependent instruments (HEIDI) tests included in the SMR software^44^. Briefly, SMR was originally developed to test for pleiotropic association between the expression level of a gene (or the methylation level of a CpG site) and a complex trait of interest using summary-level data from GWAS and QTL studies. We used the summary statistics of the aforementioned GWAS and the nominal placental *cis*-mQTL database. No significant hits were found for most traits, including early onset disorders such as ASD and ADHD, and others such as OCD or SA. In turn, multiple SMR hits (Bonferroni P_SMR_ < 0.05 and P_HEIDI_ > 0.05) were identified for BIP (n=29), MDD (n=27) and especially for SCZ (n=188) (Supplementary Data 5).

To replicate pleiotropic associations from the SMR analyses, we performed a colocalization test with the same data. The Bayesian colocalization analysis implemented in the *coloc* R package^45^ focuses on finding the intersection between significant variants independently associated with two phenotypes, comparing the association patterns in the GWAS and QTL analyses across genomic regions, and thus combining the summary statistics into posterior probabilities for five hypotheses (see Methods for more information). Colocalization was performed across 64, 102, and 287 genomic regions defined in each GWAS associated with BIP, MDD and SCZ, respectively. These regions spanned 110,721 placental DNAm sites from the nominal *cis*-mQTL database. In BIP, the posterior probabilities for 47 regions, involving 488 DNAm sites in 537 region-CpG pairs, were supportive of a colocalization signal (PPA4>0.8) (Supplementary Data 6). In MDD, the posterior probabilities for 52 regions, involving 284 DNAm sites in 295 region-CpG pairs, were supportive of a colocalization signal (PPA4>0.8) (Supplementary Data 6). Finally, in SCZ, the posterior probabilities for 188 regions, involving 2,057 DNAm sites in 2,177 region-CpG pairs, were supportive of a colocalization signal (PPA4>0.8) (Supplementary Data 6). When the overlap with SMR hits was assessed, out of the 29 SMR hits in BIP, 22 DNAm sites (75%) showed evidence of colocalization with BIP. In MDD, from the 27 SMR hits, 13 DNAm sites (48%) showed evidence of colocalization with MDD. And, from the 188 SMR hits in SCZ, 96 DNAm sites (51%) showed evidence of colocalization with SCZ.

To provide more insights into the functional relevance of our findings, we examined associations between placental DNAm and placental gene expression for the CpGs from the SMR and colocalization analyses. We interrogated expression quantitative trait methylation sites (eQTMs) from an independent set of 195 fetal placenta samples from the Rhode Island Child Health (RICHS) study^46^. Briefly, eQTMs were calculated using linear models in MatrixEQTL, adjusted by fetal sex, 5 gene expression PCs and Planet-estimated cell types, and considering all the genes in a 0.5 Mb window up and downstream of each CpG. Among the 29, 27 and 188 SMR-significant DNAm sites identified in BIP, MDD and SCZ, we found 2, 4 and 48 significant eQTM-genes (FDR<0.05), respectively (Supplementary Data 7), strongly supporting the placental function of the CpGs identified.

Next, we identified the intersect between SMR, colocalization and eQTM results for each trait (Supplementary Figure 6). In the case of BIP, two placental DNAm sites that colocalized with two GWAS loci on chromosomes 3 and 5 showed DNAm levels correlated with the expression of two nearby genes (*ITIH4* and *ZNF592*). For MDD, two placental DNAm sites colocalized with a single MDD-associated genomic locus on chromosome 14, with DNAm levels correlating with the placental expression of *LFRN5*. Finally, in SCZ, 28 placental DNAm sites colocalized with 11 SCZ-associated loci in chromosomes 3, 6, 7, 11, 12, 18, 19 and 22, with DNAm levels that correlated with the placental expression of 19 nearby genes (*FXR1, GLB1L3, HLA-H, IRF3, NAGA, PGBD1, PSMG3, RNF39, SLC6A16, TOB2P1, TRIM27, TXNL4A, VARS2, VPS37B, ZKSCAN4, ZNF165, ZNRD1-AS1, ZSCAN12P1* and *ZSCAN23*).

One of the overlapping hits in BIP was cg17890764 (Bonferroni P_SMR_ = 0.041) that colocalized with the BIP-associated region chr3:52214443-53175443 (PPA4 = 0.828). The DNAm levels of this CpG were negatively correlated with the placental expression of *ITIH4* (FDR P = 6.28×10^−4^, R2 = -0.302), a gene that has been highlighted in an integrative analysis of BIP GWAS and different regulatory SNP annotation datasets, and is known to participate in neuroinflammation^47^ (Supplementary Figure 7).

Regarding MDD, two independent, pleiotropically associated CpGs, cg23217097 and cg10318063 (Bonferroni P_SMR_ = 0.047 and P_SMR_ = 0.004, respectively), colocalized with the MDD-associated locus on chr14:41940872-42476274 (PPA4 = 0.977 and PPA4 = 0.974, respectively) (Figure 4). The DNAm levels of those CpGs correlated with the expression of *LRFN5* in placenta (FDR P = 6.08×10^−5^ and R^2^ = -0.342, and FDR P = 1.03×10^−13^ and R^2^ = 0.538, respectively). Remarkably, several SNPs in *LRFN5* have been found to be pleiotropically associated with both MDD and chronic pain, although the effector tissue or cell type have not been fully established^48^. On the other hand, it is also known that this gene is expressed in TB stem cells, and therefore could play a role in placenta^*49*^. Finally, the two independent CpGs identified are located in the promoter region of *LRFN5*. Altogether, these results suggest that the reported association is potentially causal, rather than pleiotropic.

**Figure 4.**
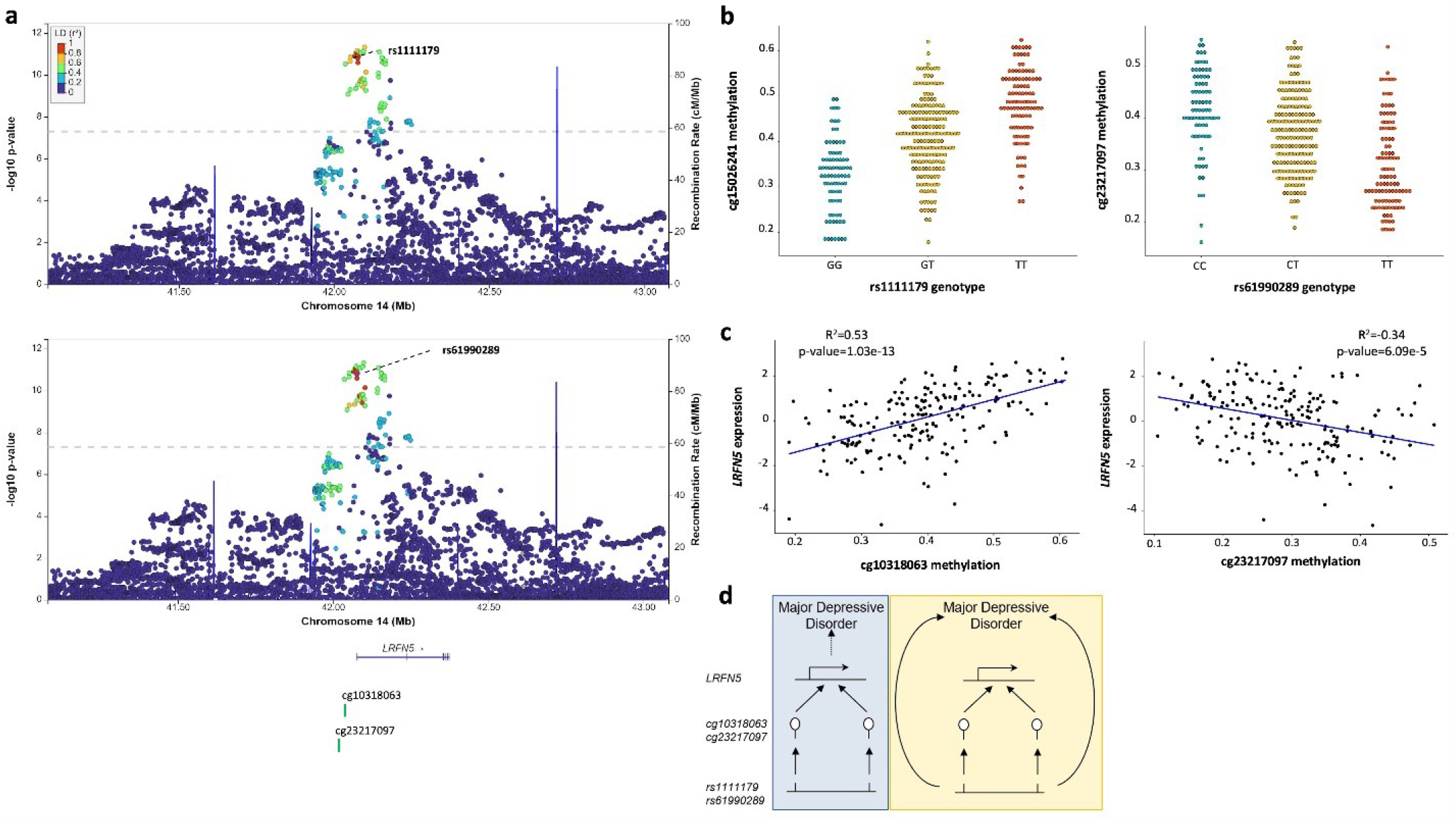
Two different CpGs pleiotropically associated with MDD. The mQTL-SNPs rs1111179 and rs61990289, highlighted as purple diamonds, with the -log_10_ p-values of the original MDD GWAS are represented in the **a** locusZoom plot. The X-axis displays the involved genomic region of chromosome 14 in Mb, showing the distribution of the coding genes in the locus, as well as the location of the cg10318063 and cg23217097 mQTL-CpGs. The Y-axis shows the -log_10_ p-value from the original GWAS, and the SNPs are colour coded as a function of the LD with the highlighted SNP. The two significant mQTLs, rs1111179-cg10318063 and rs61990289-cg32217097, are plotted in the **b** dotplots, where Y-axis represents the beta DNAm values of the indicated CpG, ranging from 0 to 1. The X-axes display the genotype of the indicated SNPs. The eQTM of cg10318063 and cg32217097 mQTL-CpGs are portrayed in the **c** dotplots, where X-axis represents the DNAm values from each involved CpG, ranging from 0 to 1. The Y-axes display the expression values of the *LFRN5* gene in placenta. The hypothesis of the pleiotropical association between the SNPs, the DNAm values of the CpGs in placenta, the gene expression levels of *LFRN5* in placenta and MDD are schematically represented in **d**. The vertical pleiotropy (or causal association) hypothesis is represented with a blue backgroud, and the horizontal pleiotropy hypothesis is highlighted with a yellow background.

In SCZ, cg03172226 was found to be pleiotropically associated with the disorder (Bonferroni P_SMR_ = 0.032), colocalized with the GWAS peak on chr19:50007909-50252109 (PPA4 = 0.999), and DNAm levels of the CpG were correlated with placental expression of *IRF3* (FDR P = 0.018 and R^2^ = 0.237) (Supplementary Figure 8). Importantly, *IRF3* is a regulator of type I interferons with a pivotal role in MIA^50^. Another example of pleiotropic association with SCZ is cg17805547 (Bonferroni P_SMR_ = 0.032). This CpG colocalized with the SCZ-associated regions chr3:179745069-180418069 and chr3:180475148-181295605 (PPA4 = 0.864 and 0.862, respectively), and its DNAm levels were correlated with the expression of *FXR1* in placenta (FDR P = 0.027 and R^2^ = 0.227) (Supplementary Figure 9). *FXR1* promotes TB migration and its mRNA levels have been reported to be decreased in TB from women with recurrent spontaneous abortions^51^. Additionally, *FXR1* regulation of interneurons in the prefrontal cortex is critical for SCZ-like behaviors^52^.

Finally, we performed a Reactome gene-set analysis^53^ of the 19 genes at the intersection of the three approaches in SCZ. We observed an enrichment for 10 gene-sets (Supplementary Data 8), including immune system pathways such as interferon alpha/beta signaling, interferon gamma signaling, and interferon signaling, supporting the idea of MIA as a link between placenta and SCZ risk.

### Conditional SMR analysis of placental *cis*-mQTLs, and BIP, MDD and SCZ

The developers of the SMR software designed the HEIDI test assuming a single causal variant in the *cis*-mQTL region that affects both DNAm and the trait analyzed^44^. Under the assumption of pleiotropy, when there are multiple causal variants in a region, the pleiotropic signal of one causal variant will be diluted by that of other non-pleiotropic causal variants. We therefore performed a GCTA conditional analysis^54^ conditioning for the top associated *cis*-mQTL in both the GWAS and mQTL data sets. If there was a secondary signal, pointed by the presence of heterogeneity (P_HEIDI_ < 0.05) in the *cis*-mQTL region, in either the GWAS or the mQTL dataset, we performed another round of conditional analyses conditioning only on the secondary signal in both the GWAS and mQTL datasets, and then reran the SMR and HEIDI test at the top *cis*-mQTL using the estimates of SNP effects from the conditional analyses. We applied this approach to those CpGs that passed the SMR test but failed to pass the HEIDI test. Nevertheless, due to power constrains, only those CpGs that colocalized with one of the three studied disorders according to *coloc* were considered.

We discovered a secondary hit in SCZ, in which two independently associated SNPs, i.e. rs3132386 (the primary signal) and rs16897420 (the secondary signal) had a pleiotropic effect on both cg15026241 and SCZ (Bonferroni P_SMR_ = 0.022 and P_SMR_ = 1.67×10^−4^, respectively) (Supplementary Data 5 and 9). This CpG, located in the HLA region, is correlated with the expression of *ZSCAN23* in fetal placenta (FDR P = 0.002 and R^2^ = -0.282) (Figure 5 and Supplementary Data 7). A strictly intergenic transcript downstream of this gene has already been identified in another SCZ study using a conditional GWAS/eQTL analysis, but it has not been characterized in detail^55^. Our results, namely the presence of two independent signals pointing to a single DNAm site that seems to affect gene expression in fetal placenta, together with the fact that the p-values of those SNPs do not reach genome-wide significance in the original SCZ-GWAS, strongly support causality.

**Figure 5.**
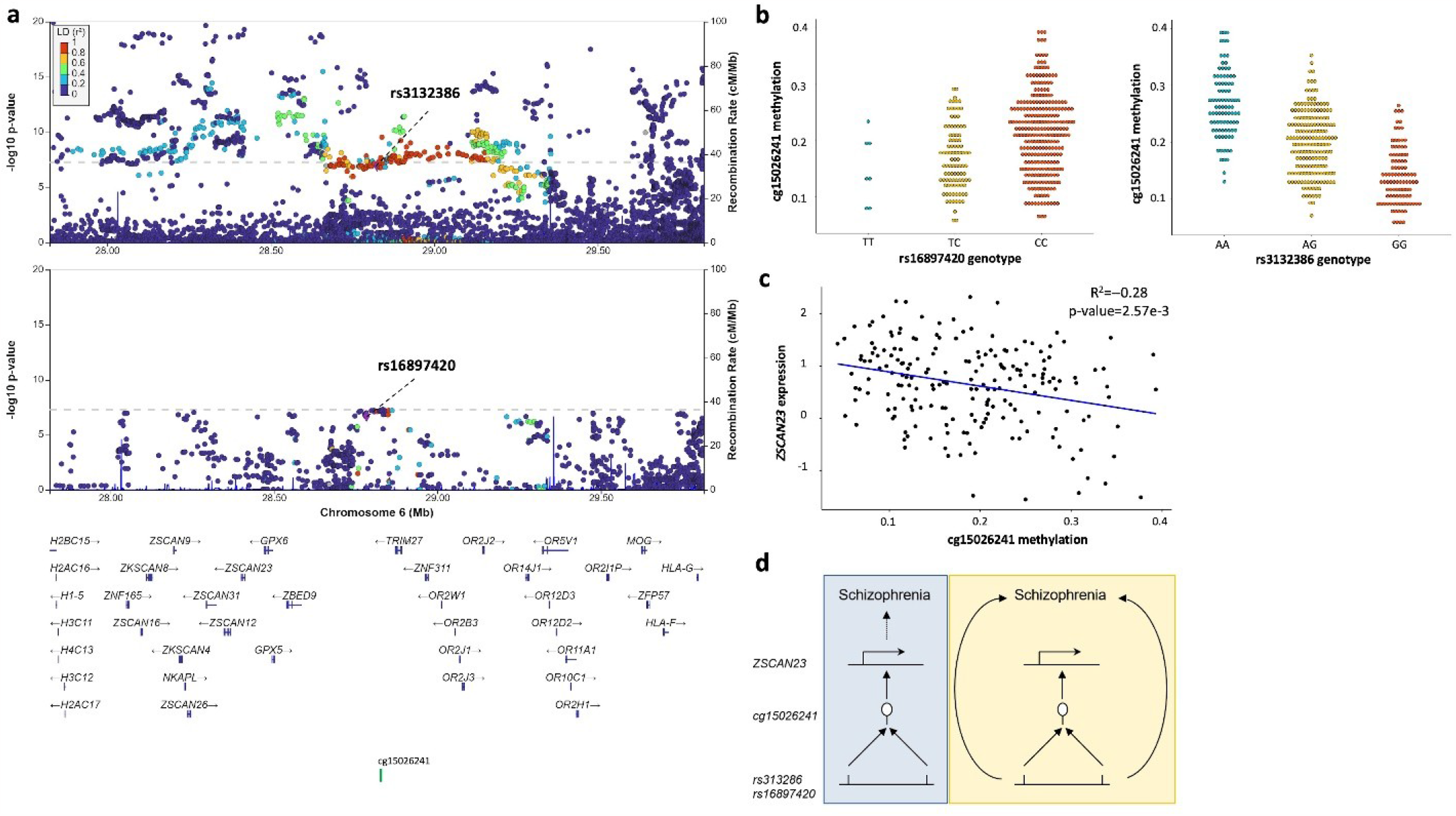
One CpGs is pleiotropically associated to SCZ, and marked by two independent signals. The two mQTL-SNPs rs313286 and rs16897420, highlighted as purple diamonds, are shown in the original (top) and the conditional (bottom) SCZ GWAS in the **a** locusZoom plot. The X-axis displays the involved genomic region in chromosome 6 in Mb, showing the distribution of the coding genes in the locus, as well as the location of the mQTL-CpG cg15026241. The Y-axis shows the -log_10_ p-value from the original and conditional GWAS, and the SNPs are colour coded as a function of the LD with the highlighted SNP. The two significant mQTLs, rs16897420-cg15026241 and rs3132386-cg15026241, are plotted in the **b** dotplots, where Y-axes represent the cg15026241 beta DNAm values, ranging from 0 to 1. The X-axis displays the genotype of the corresponding SNPs. The eQTM of the significant mQTL-CpG is portrayed in the **c** dotplot, where the X-axis shows the DNAm values of the cg15026241 CpG, ranging from 0 to 1. The Y-axis displays the expression values of the *ZSCAN23* gene in placenta. The hypothesis of the pleiotropical association between the SNPs, the DNAm values of the CpGs in placenta, the gene expression levels of *ZSCAN23* in placenta and MDD are schematically represented in **d**. The vertical pleiotropy (or causal association) hypothesis is represented with a blue backgroud, and the horizontal pleiotropy hypothesis is highlighted with a yellow background.

### SMR analysis of placental cell type- and GA-imQTLs in BIP, MDD and SCZ

We used SMR to combine imQTLs with the BIP, MDD and SCZ GWAS, and obtained two STB-imQTLs (one each in BIP and SCZ), five TB-imQTLs (two in BIP, one in MDD and two in SCZ) and two GA-imQTLs (one each in BIP and SCZ) (Supplementary Data 10 and Supplementary Figure 10). Remarkably, we found an imQTL involving cg27130493 and rs72743436 (Figure 6). cg27130493 appeared to be pleiotropically associated with BIP while it changed its methylation levels as a function of STB proportion, in a rs72743436 genotype-dependent manner. This CpG is located on the gene body of *SMAD3*. The placental overexpression of this gene has been described to activate the ability of TB to form endothelial-like networks, while its defect has been associated with pre-eclampsia^56^. Additionally, it is a well-known target for lithium treatment in BIP^57^. Finally, rs72743436 is not directly associated with BIP in the original GWAS, suggesting a possible causal association between placental DNAm and BIP at this CpG site. Another interesting imQTL-CpG was cg04402684, located on a CTCF binding region, 32 kb upstream of the *HLA-E* gene. This CpG was significant in the SMR analyses performed with TB-imQTLs in both BIP and SCZ, revealing a common DNAm site associated to both disorders (Supplementary Figure 10).

**Figure 6.**
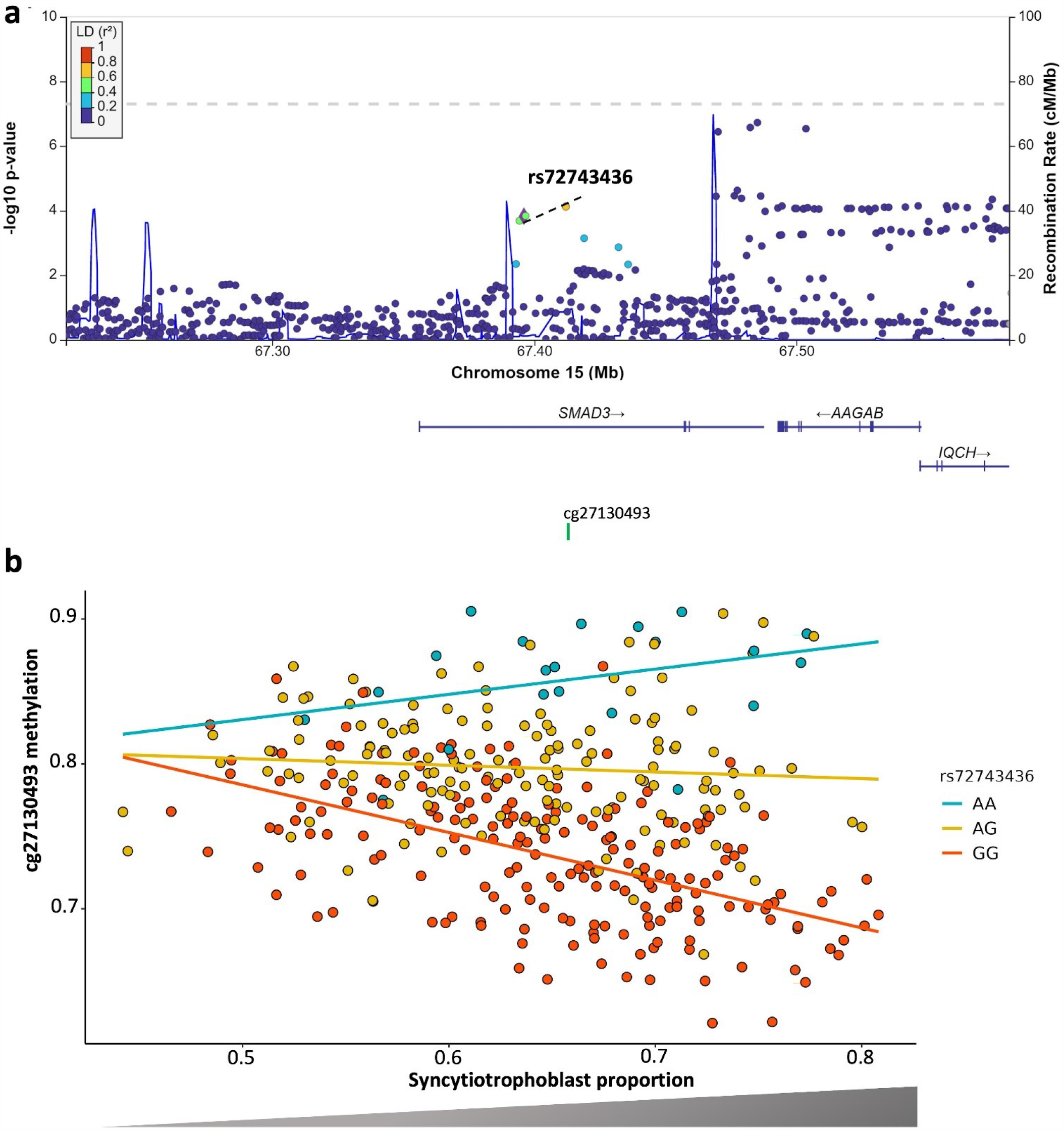
STB-imQTL pleiotropically associated with BIP. The mQTL-SNP rs72743436, highlighted as a purple diamond, is shown in the original BIP GWAS in the **a** locusZoom plot. The X-axis displays the involved genomic region in chromosome 15 in Mb, showing the distribution of the coding genes in the locus, as well as the location of the mQTL-CpG cg27130493. The Y-axis shows the -log_10_ p-value from the original GWAS, and the SNPs are colour coded as a function of LD with the highlighted SNP. The STB-imQTL is pictured in the **b** dotplot. The X-axis represents the cg27130493 beta DNAm values and the Y-axis the STB proportion, both ranging from 0 to 1. The genotype of rs72743436 SNP-imQTL is colour coded as indicated in the legend.

### Comparison with SMR results for brain mQTLs

Finally, we aimed to ascertain the tissue specificity of our findings, and thus we performed the abovementioned SMR analyses with fetal brain and brain *cis*-mQTLs in BIP, MDD and SCZ. The fetal brain *cis*-mQTL database was published by Hannon et al. in 2015^58^. Briefly, mQTLs were calculated in 166 human fetal brain samples (56-166 days post-conception). The methylation data was obtained with the Infinium HumanMethylation 450K array from Illumina, and the mapping of the mQTLs was performed with the MatrixEQTL R package^59^, considering a *cis*-window of 0.05 Mb. Only mQTLs with a P < 1.5×10^−9^ were made available, resulting in 556,513 fetal brain *cis*-mQTLs that were downloaded from the SMR portal. The second brain *cis*-mQTL database was originally from a 2018 publication by Qi et al.^60^, in which *cis*-mQTLs from three different datasets were meta-analyzed: in particular, 468 brain cortical region samples from the ROSMAP study^61^, 166 fetal brain samples by Hannon et al., and 526 frontal cortex region samples^62^ were included, amounting to a final sample size of 1,160 brain samples and nearly 6M *cis*-mQTLs. As in the case of the fetal brain mQTL database, this dataset was downloaded from the SMR data portal.

In BIP we obtained 13 and 35 significant SMR hits (Bonferroni P_SMR_ < 0.05 and P_HEIDI_ > 0.05) in the fetal brain and brain mQTL datasets, respectively (Supplementary Data 11 and Supplementary Figure 11), and with same criteria, we obtained 10 and 23 SMR hits in MDD, and 50 and 188 in SCZ. As shown in Figure 7, the trait with the largest overlap of pleiotropically associated DNAm sites among tissues is SCZ, although the overlap is very limited in all the traits. In the case of the fetal brain database, the smaller sample size limits the number of detected mQTLs and could lead to underestimate the proportion of the genetic risk that acts through the fetal brain. When we compared our most reliable hits in placenta (intersection among SMR, *coloc* and eQTM results) with the brain SMR results, neither BIP or MDD presented any overlap of DNAm sites, while there were two common hits for SCZ. cg03172226, the placental eQTM for *IRF3*, was also identified in the brain dataset in combination with rs7251, the same SNP identified in placenta. On the other hand, cg21745287, the placental eQTM for *VPS37B*, overlaps with the fetal brain results, although in combination with a different SNP than in placenta. Therefore, we cannot rule out that mostly in the case of these non-specific shared DNAm sites, the placenta could be mirroring what occurs in the brain without necessarily being the main effector tissue.

**Figure 7.**
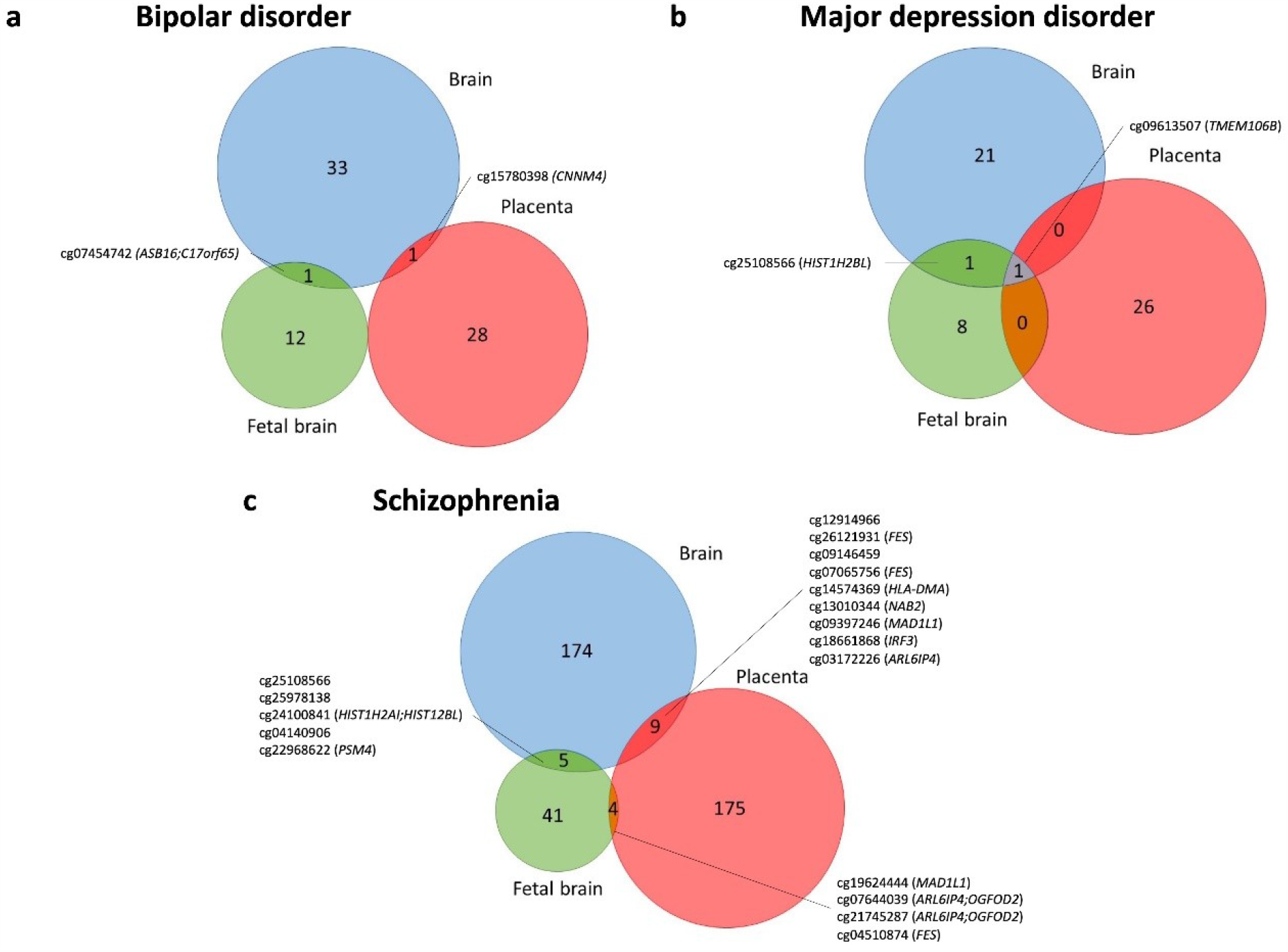
Overlap between the SMR results in brain, placenta and fetal brain, in BIP, MDD and SCZ. The overlap between the mQTL-CpGs pleiotropically associated between the three tissues and BIP, MDD and SCZ are represented in the **a, b** and **c** Venn diagrams, respectively. Overlapping mQTL-CpGs are shown, and also the closest gene from the Illumina annotation file (between brackets).

## Discussion

In this study, we have constructed three different placental *cis*-mQTL databases in 368 placenta samples from the INMA project: the nominal placental *cis*-mQTL database, in which all SNP-CpG combinations over a nominal p-value cutoff of 5×10^−8^ in 0.5 Mb windows have been compiled; the permuted placental *cis*-mQTL database, in which we have corrected for multiple correlated variants considering all the possible combinations in the same windows as above, and the conditional placental *cis*-mQTL database, that uses the permuted mQTLs to perform a stepwise regression procedure that enables to conditionally map independent *cis*-mQTLs. Importantly, we have made all results publicly available both in their raw formats and by means of a user-friendly shiny-based genome browser that enables to search for the mQTLs of interest by CpG, SNP and/or genomic coordinates. We believe that this tool will be useful to the scientific community.

In general, the placental *cis*-mQTLs in all three databases were depleted in regions that are usually hypomethylated, such as promoters and CpG islands, very likely due to the fact that lower and more stable DNAm values will hardly correlate with the genotype or any other variable. In turn, an enrichment of intermediate DNAm values was observed, usually present in CpG island shelves and shores, as well as in open sea regions. The genotype-regulated placental DNAm seems highly placenta-specific and interestingly, is enriched in placenta-specific active chromatin marks, and in developmental disorders of mental health and psychotic disorders, including SCZ. This, together with the fact that mQTLs have recently been highlighted as powerful instruments to reveal molecular links to traits otherwise missed by eQTL-GWAS colocalization approaches^22^, encouraged us to conduct a multi-omics study to try to unveil the placental contribution on the developmental origins of different neuropsychiatric disorders.

Besides, Kim-Hellmuth et al. described cell type-interacting expression QTLs (ieQTLs) to be enriched in GWAS signals and to improve GWAS-eQTL matching for the mechanistic understanding of these loci^23^. In this context, we wondered whether this could also be true for placental mQTLs and calculated STB- and TB-imQTLs. STB were considered for several reasons, 1) they are the most abundant cell type in term placenta^63^, 2) they cover the entire surface of villous trees in the placenta, and thus are in direct contact with maternal blood^64^, 3) they orchestrate the complex biomolecular interactions between the fetus and the mother, and 4) they act as an important endocrine organ, producing numerous growth factors and hormones that support and regulate placental and fetal development and growth^65,66,67,68^. Given that TB are STB progenitor cells, and that there is a very high correlation between the proportions of these two cell types in our samples, we decided to calculate TB-imQTLs as well.

We observed higher statistical power for the most abundant cell type and therefore, a larger amount of STB-imQTLs compared to TB-imQTLs. However, there was a considerable overlap and 46.3% of STB-imQTLs were also TB-imQTLs. As expected, the allelic effects in the overlapping imQTLs were negatively correlated between the two cell type-specific models, with imQTLs positively correlated with STB content being more common. This could reflect that negative interactions with a given cell type more likely reflect interactions with another cell population that is decreasing, rather than a truly negative, genotype-dependent correlation. As TB are known to differentiate into STB throughout the gestation^30^, and STB content in term placenta is positively correlated with GA at birth, we wanted to know whether cell type-imQTLs are equivalent to GA-imQTLs. This was not the case, revealing that GA has an independent effect on placental DNAm other than the TB to STB differentiation.

Remarkably, placental DNAm is pleiotropically associated with BIP, MDD, and in particular with SCZ, while it does not seem to associate with early onset conditions such as ADHD and ASD. These results could arise from the sample sizes and power constraints of the available GWAS, as well as, from the higher heritability of certain disorders, including BIP and SCZ. However, very importantly, a recent article found out that MDD shows a very high polygenicity compared to other psychiatric disorders; that is, more genetic variants with weaker effects contribute to the overall genetic signal in MDD, and make the trait less annotatable^69^. In contrast, ADHD, BIP and SCZ showed the highest discoverability and hence a more annotatable genetic signal. Subsequently, the estimated sample size required to reach 90% SNP heritability was more than eight times larger for MDD than for ADHD, BIP and SCZ. Therefore, the lack of signal in ADHD and even more remarkably, the presence of a considerable pleiotropy between placental DNAm and MDD, suggest that our findings are guided not only by the strength and annotatability of the genetic basis of the diseases studied, but rather by a genuine association with placental DNAm. In addition, it is well known that BIP, MDD and SCZ share a common genetic background and therefore, it is plausible that part of the genetic risk could act through common processes at similar developmental stages^70^.

It is possible to gain insight into the pathogenesis of complex disorders by defining the environmental, biological and temporal context in which genes increase disease susceptibility. In 2018, Ursini et al discovered that when early-life complications are present, the polygenic risk score (PRS)-explained risk of SCZ is more than five times higher than when they are absent^71^. SCZ loci that interact with early-life complications are not only highly expressed in placenta, but also differentially expressed between complicated and normal pregnancies in this organ, and enriched in response-to-stress pathways. More recently, these placenta-specific SCZ-PRS have also been shown to interact with both brain volume and cognitive function, suggesting particular neurodevelopmental trajectories in the path towards SCZ^72^. Particularly in the latter work, they also studied the existence of placenta-specific PRS in other neuropsychiatric disorders, and if present, whether they interacted with early neurodevelopmental outcomes. BIP presented a PRS with a very high enrichment in genes that are expressed in placenta, although this placenta-specific PRS did not interact with neither brain volume nor cognition. This makes sense since SCZ and BIP present the highest genetic correlation compared to any other pair of psychiatric traits^73^, but with remarkable differences in outcome, due to the fact that SCZ patients have been described to be more prone to suffer from severe cognitive impairment than BIP patients^74^. In conclusion, placental DNAm could be important in BIP although through trajectories that are different from those characterized by Ursini and colleagues in SCZ. In any case, we believe that our findings support a novel hypothesis according to which placental DNAm could translate both the genetic basis and the environmental milieu into fetal genetic programs that could eventually result in impaired neurodevelopmental trajectories leading to SCZ, and maybe, to other neuropsychiatric disorders. This hypothesis will need to be supplemented with additional research.

Regarding the genes and pathways affected by placental DNAm pleiotropically associated to SCZ, we want to highlight that the 19 genes resulting from our multi-omics approach were enriched in immune-related terms, including interferon signaling. *HLA-H, IRF3* and *VPS37B* were some of the most relevant genes with immune functions. This reinforces the idea of MIA being implicated in the neurodevelopmental origins of SCZ. During pregnancy, environmental insults such as early-life complications and maternal infections, are hypothesized to program the immune and developmental epigenetic code in the fetus, thereby influencing the risk to suffer from neurodevelopmental disorders later in life^75,76,77^. The MIA hypothesis proposes that exposure to a dysregulated maternal immune milieu *in utero* affects fetal neurodevelopment^78,79^. Moreover, in humans, maternal factors implicated in MIA are associated with epigenetic modifications in placenta^80,81^. The placenta plays a pivotal role in maintaining immune homeostasis in the maternal-fetal interface. However, when a sustained placental inflammatory response occurs due to maternal environmental factors, the offspring can suffer from developmental abnormalities^82^.

Taken individually, the hits that are most likely to be causally involved in the different neuropsychiatric disorders studied were *LRFN5* in MDD and *ZSCAN23* in SCZ. In the case of *LRFN5*, several variants in this gene have been found to be pleiotropically associated with both MDD and chronic pain^48^. Additionally, it is well known that *LRFN5* is involved in the communication among brain cells and is located in a large and complex genomic niche that is highly conserved in mammals^83^. Specific locus structure of this region increases ASD susceptibility in males. But what makes *LRFN5* especially notable in the context of our study is the fact that we have found regional pleiotropy, that is, two independently associated CpGs in its promoter that are correlated with its expression levels in fetal placenta. These two independent signals, together with the DNAm-expression correlation in our organ of interest make the association more likely causal.

In the case of *ZSCAN23*, in a work published in 2019, conditional analyses were performed to partition the SCZ GWAS-associated brain eQTLs to better identify the downstream molecular features of genetic risk^55^. The top GWAS risk variant rs1233578 was associated with a strictly intergenic sequence downstream of *ZSCAN23*, and the authors claimed to have discovered a novel transcript potentially involved in SCZ through its expression in brain. This is not necessarily contradictory with our results. The function of *ZSCAN23* is unknown, although a recent paper has reported that two CpGs in the gene could show differentially variable methylation in placenta between asthmatic and control mothers^84^. In the present work, two SNPs have been found to independently associate with the same CpG that is related to the expression of *ZSCAN23* in placenta. Moreover, the GWAS p-values of the association between the two SNPs and SCZ are not significant. This makes placental DNAm and *ZSCAN23* expression potentially causal in SCZ, although the function of the gene and the pathways through which it may act remain unknown.

In this context, the placental DNAm sites that resulted from the SMR analyses confronting the BIP, MDD and SCZ GWAS, and the STB-, TB-, and GA-imQTLs are worthy of mention. For example, we found out that cg27130493 is pleiotropically associated with BIP while it changes its methylation levels as a function of STB proportion, in a rs72743436 genotype-dependent manner. This CpG is located in the gene body of *SMAD3*, a gene with singular functions in placenta and also in BIP itself^56,57^. Additionally, this SNP is not directly associated with BIP, increasing the likelihood of causality rather than pleiotropy. The most important limitation of this part of the study is the lack of placental cell type-specific expression data to ascertain whether cg27130493 and other DNAm sites identified in this approach are correlated with the expression of nearby genes in placenta. However, the fact that the CpG sites change not only depending on the genotype of adjacent SNPs, but also as a function of estimated placenta-specific cell content, increases the probability of placenta being the effector organ of those genetic associations.

Lastly, we tried to ascertain the tissue-specificity of our findings by comparing placenta SMR hits to those from two brain *cis*-mQTL databases. We found a limited overlap among the different tissues studied, suggesting that the majority of our pleiotropically associated DNAm sites could be relatively specific of placenta (or at least, not related to brain DNAm). Nevertheless, *IRF3*, one of the most promising candidates due to its immune function, overlaps with the brain database. We cannot rule out neither that placenta is mirroring the brain with no effector role, nor that *IRF3* has a role in SCZ through different tissues, organs and developmental stages. Finally, it is important to consider that, although the SMR approach was executed exactly in the same manner for the three databases, these were quite different from each other. Especially the fetal brain database was calculated in a limited number of fetal brain samples, in SNP-CpG windows 10 times smaller than the ones we used (0.05 vs 0.5 Mb), and DNAm was measured with the Infinium HumanMethylation 450K array (with approximately half the probes in the Infinium HumanMethylation EPIC array used in the present study), therefore resulting in half a million *cis*-mQTLs compared to our nearly 8 million. This suggests a more limited mapping potential with the fetal brain mQTL database and thus, a possible underestimation of the fetal brain DNAm that is really involved in the disorders studied.

The main limitation of our study is that, as pointed out by Ursini et al in relation to their own work^72^, the considerable overlap in cell biology between brain and placenta does not allow to exclude the possibility that part of the pleiotropy observed here is related to a more direct effect in the brain exerted by the same DNAm sites as in placenta. However, we believe that the different pieces of evidence, including the intersection with the placental eQTMs and even the presence of secondary signals, support that a part of the genetic risk to suffer from BIP, MDD and very especially SCZ, could act through placental DNAm at specific genomic loci. Another limitation is that different methylation beadchips have been employed in the INMA and RICHS cohorts, and thus, we are probably underestimating the effects of DNAm over placental gene expression.

In conclusion, we find placental *cis*-mQTLs to be highly placenta-specific, with a remarkable enrichment in genomic regions active in placenta and neurodevelopment- and mental health-related pathways. We prove that they are useful to map the etiologic window of neuropsychiatric disorders to prenatal stages and conclude that part of the genetic risk of BIP, MDD and in particular SCZ, act through placental DNAm at specific genomic loci. In fact, some of the observed associations might be causal rather than pleiotropic due to the presence of secondary association signals in conditional analyses, regional pleiotropic DNAm associated to the same disorder, and involvement of cell type-specific imQTLs, that additionally associate to the expression levels of relevant genes in placenta. It is of particular interest that SCZ-associated placental DNAm correlates with the expression of immune-related genes in placenta, providing further support to the hypothesis of the neurodevelopmental origins of SCZ.

## Methods

### Placental biopsies and DNA extraction

In the INMA^24^ project, 2,506 mother-fetus pairs were followed until birth and a selection of 397 placentas were collected, representing the three geographical areas involved in the study. Collected placentas were stored at -80°C in a central biobank until processing. Biopsies of approximately 5 cm^3^ were obtained from the inner region of the placenta, approximately 1.0-1.5 cm below the fetal membranes, corresponding to the villous parenchyma, and at a distance of ∼5 cm from site of cord insertion. 25 mg of placental tissue was used for DNA extraction, previously rinsed twice during 5 minutes in 0.8 mL of 0.5X PBS in order to remove traces of maternal blood. Genomic DNA from placenta was isolated using the DNAeasy® Blood and Tissue Kit (Qiagen, CA, USA). DNA quality was evaluated on a NanoDrop spectrophotomer (Thermo Scientific, Waltham, MA, USA) and additionally, 100 ng of DNA was run on 1.3% agarose gels to confirm that samples did not present visual signs of degradation. Isolated genomic DNA was stored at -20°C until further processing.

### Genotype data

Genome-wide genotyping was performed using the Illumina GSA Beadchip at the Human Genotyping Facility (HuGeF), Dept Internal Medicine, Erasmus MC, Rotterdam, the Netherlands, and the Spanish National Genotyping Centre, CEGEN, Madrid, Spain. Genotype calling was done using the GeneTrain2.0 algorithm based on HapMap clusters implemented in the GenomeStudio software. Samples were genotyped in four batches.

The quality control of the genotype data from 397 INMA samples and 509,450 genetic variants was performed using the PLINK 1.9 software following the standard recommendations^85,86,87^. All plink files were initially processed with Will Rayner’s preparation Perl script available from Mark McCarthy’s Group as recommended in the documentation from the Michigan Imputation Server, using the Haplotype Reference Consortium (HRC) r1.1 2016 reference panel^88^. Variants with a call rate below 95%, minor allele frequency (MAF) below 1%, or a p-value from the Hardy-Weinberg Equilibrium (HWE) exact test below 1×10^−6^ were removed. Samples with discordant sex, those with average heterozygosity values above or below 4 standard deviations or with more than 3% missing genotype were filtered out. Identity-by-descent values were calculated with PLINK, and from those sample pairs that showed PI-HAT estimates above 0.18, the sample with higher proportion of missing genotypes was removed.

The final dataset was imputed with the Michigan Imputation Server^88^ using the HRC reference panel, Version r.1.1 2016^89^. Before imputation, data was converted into VCF format. Phasing of haplotypes was done with Eagle v2.4^90^ and genotype imputation with Minimac4^91^, both implemented in the code by the Michigan Imputation Server. Finally, we removed variants with an imputation dosage r^2^ below 0.9, a MAF lower than 5%, a HWE p-value below 0.05 and with more than two alleles, to avoid SNPs with few or no individuals bearing the minor allele homozygous genotype in our sample set. Only those samples with paired methylation data were considered in this analysis. The final dataset consisted in 368 samples and 4,171,035 SNPs.

### Methylation data

DNA methylation was assessed with the Infinium MethylationEPIC BeadChip from Illumina, following manufacturer’s protocol in the Erasmus Medical Centre core facility. Briefly, 750 ng of DNA from 397 placental samples were bisulphite-converted using the EZ 96-DNA methylation kit from Zymo Research, following the manufacturer’s standard protocol, and DNA methylation was measured using the Infinium protocol. Three technical duplicates were included. Samples were randomized taking into account region-of-origin and sex. As the number of samples in each condition was different, a perfect randomization was not possible. However, all the plates had samples from all three involved geographical areas, and an equilibrated number of male and female samples.

The quality control of the methylation data, including 865,859 DNAm probes, was performed using the PACEAnalysis R package (v.0.1.7)^92^. Before starting the quality control with PACEAnalysis, one sample was discarded for having too many missing values in relevant variables. With the R package, we discarded those samples with a call rate below 95%, sex inconsistencies, intentioned or non-intentioned duplicates and exhibiting contamination with DNA from another subject or the mother. Only those samples with paired genotype data were considered in this study. Probes with a call rate smaller than 95%, within sexual chromosomes, SNPs (European MAF < 5%) and cross-hybridizing potential were excluded from the analysis.

The methylation beta values were normalized in different steps. Dye-bias and Noob background correction, implemented in minfi R package, were applied^93,94^, followed by normalization of the data with the functional normalization method^95^. Then, to correct for the bias of type-2 probes values the beta-mixture quantile (BMIQ) normalization was applied^96^. After that, we explored the clustering of the data through Principal Component Analysis (PCA) and tested the association of the 12 first PCs with the main and the technical variables. Array batch effect was controlled with the ComBat method^97^. Finally, to correct for the possible outliers, we winzorized the extreme values to the 1% percentile (0.5% in each side), where percentiles were estimated with the empirical beta-distribution. The final dataset consisted in 368 samples and 747,486 DNAm probes (CpGs).

Cell type proportions of six populations (STB, TB, nucleated red blood cells, Hofbauer cells, endothelial cells, and stromal cells) were estimated from DNAm using the placenta reference panel from the 3^rd^ trimester implemented in the Planet R package^31^.

### Placental *cis-*mQTL analysis

A total amount of 4,171,035 SNPs, 747,486 CpGs and 368 samples with paired genotype and DNAm data were considered for the *cis*-mQTL analysis in TensorQTL^98^. TensorQTL nominal modality performs linear regressions between the genotype and the normalized DNAm beta-values, as implemented in FastQTL^99^. The covariates included in the regression model were the sex of the fetuses, the first five PCs derived from the genotype data (genotype PCs) and the five cell type proportions derived from the Planet methylation panel. Genotype PCs were included in the model as covariates to remove the hidden batch effects and other potential confounders in the genotype data. The TensorQTL *cis*-region was specified as ±0.5 Mb from each tested CpG position, consistent with the results of previous studies where the distance between the majority *cis*-mQTL SNPs and DNAm sites is ≤ 0.5 Mb^59,100,101,102^. In the final nominal *cis*-mQTL database we included only those probes with at least one *cis*-mQTL at P_nominal_ < 5×10^−8^. This same regression model was used to build two additional *cis*-mQTL databases; permuted and conditional. The permuted *cis*-mQTL database consists in correcting for multiple correlated variants tested via a beta approximation which models the permutation outcome with a beta distribution as it is described in Ongen et al. 2016^99^. Conversely, the conditional database uses the permuted QTLs to perform a stepwise regression procedure to map conditionally independent *cis*-QTLs as described by the GTEx Consortium^103^.

### Characterization of placental *cis*-mQTLs

The separate characterization of the nominal, permuted and conditional mQTL databases was performed in different steps. First, the distribution of both, the distance between mQTL-CpGs and paired mQTL-SNPs, and the mQTLs along the chromosomes was plotted, followed by a volcano plot to check the uniform distribution of the negative and positive effect sizes.

Second, using the annotation from IlluminaHumanMethylationEPICanno.ilm10b4.hg19 R package^104^, we performed several Chi-Square tests to assess the enrichment and depletion of the UCSC RefGene and Relation to CpG Island annotations. We also depicted density plots of the methylation values according to the Relation to CpG Island annotation. Moreover, for the top 10,000 mQTL-CpGs, we assessed enrichment and depletion of overlap with tissue-specific and cell-type-specific regulatory features including DNase I hypersensitivity sites (DHS), all 15-state chromatin marks, and all five H3 histone marks (i.e., H3K27me3, H3K4me1, H3K4me3, H3K36me3, H3K9me3) from consolidated ROADMAP Epigenomics Mapping Consortium^105^ using eFORGE v2.0^26,27,28^. The enrichment and depletion with each of the three putative functional elements were tested separately and compared to the respective data from the consolidated ROADMAP epigenomics reference panel.

Lastly, over-representation analyses of Gene Ontology (GO) and Kyoto Encyclopaedia of Genes and Genomes (KEGG) gene sets were conducted with MissMethyl R package^106,107,108,109^. MissMethyl performs a hypergeometric test taking into account the bias derived from the differing number of probes per gene and the multiple genes annotated per CpG^106^. Gene set enrichment analyses with the Disease Ontology (DO) database were conducted with the DOSE R package^110^. With DOSE, we performed the gene set enrichment analysis considering the genes annotated in the IllumuminaHumanMethylationEPICanno.ilm10b4.hg19 R package^104^ as background.

### Placental *cis*-imQTL analysis

The cell type-imQTLs were computed with the interaction modality from TensorQTL, consisting in nominal associations for a linear model that includes a genotype per interaction term^98^. The same initial SNPs, CpGs and samples from the nominal database were considered for the interaction analysis. The covariates used were the sex of the fetuses and the first five genotype PCs. Following the recommendations by Kim-Hellmuth S. et al.2021^23^, estimations of STBs and TBs per sample were defined separately as the interaction terms. In the two final interaction *cis*-mQTL databases, we included only probes with at least one *cis*-mQTL at P_nominal_ < 5×10^−8^. The imQTLs obtained from each model were categorized as positively and negatively correlated with cell type estimates, or as uncertain if this was ambiguous. To categorize the imQTLs into these groups, genotype main effects at low (<25^th^ percentile) vs high (>75^th^ percentile) cell type proportion were compared. imQTLs with positive cell type correlation showed an increase of the genotype main effect from low to high cell proportions (*β*_*mQTL*_ low < *β*_*mQTL*_ *high*). imQTLs with negative cell type correlation showed a decrease (*β*_*mQTL*_ low > *β*_*mQTL*_ *high*) and the uncertain group contained imQTLs where the sign flipped between low and high cell type proportions (*β*_*mQTL*_ low ≠ *β*_*mQTL*_ *high*).

Additionally, GA was also considered as interaction term. In this case, the covariates considered were the sex of the fetuses, the first five genotype PCs and the cell type proportions from Planet.

### Genome Wide Association Studies

The GWAS summary-statistics used in this analysis were the public largest to-date association studies of ADHD^32^, AGR^33^, ASD^34^, BIP^35^, INT^36^, MDD^37^, OCD^38^, PD^39^, SA^40^, and SCZ^41^ from EAGLE, Indonesia Schizophrenia Consortium, International Obsessive Compulsive Disorder Foundation Genetics Collaborative (IOCDF-GC), OCD Collaborative Genetics Association Studies (OCGAS), PsychENCODE, Psychiatric Genomics Consortium (PGC), Psychosis Endophenotypes International Consortium, UK Biobank (UKB), SynGO Consortium, and 23andMe, among others.

All the GWAS summary-statistics were harmonized according to the dbSNP build 155 and the INMA genotype data as a reference. The harmonization steps included: to change rsID to chromosome:position nomenclatures, to correct the effect allele, the effect size and the allele frequency according to the reference genotypes if applicable, and to create a .ma format file with the summary-statistics as indicated in the SMR pipeline^44^.

### Multi-SNP-based Mendelian Randomization analysis

Mendelian Randomization analysis was carried out considering *cis*-mQTL SNPs as the instrumental variables (IVs), CpG methylation as the exposure (X), and the neuropsychiatric traits as the outcome (Y). Multi-SNP-based MR (SMR-multi)^111^ analysis was performed by the SMR software^43^. SMR integrates GWAS and QTL summary-statistics to test for pleiotropic associations between quantitative traits, such as methylation or expression, and a complex trait, for instance a disease. More precisely, SMR-multi includes multiple SNPs at a *cis*-mQTL locus in the SMR test to calculate the causative effect of an exposure on an outcome (*b*_*xy*_). First, SMR-multi selected all SNPs with P_nominal_ < 5×10^−8^ in the *cis* region (0.5 Mb of the CpG). Second, it removed SNPs in very high LD with the top associated SNP (LD r^2^ > 0.9). Then, the causative effect (*b*_*xy*_) from the exposure on the outcome was estimated at each of the SNPs and combined in a single test using an approximate set-based test accounting for LD among SNPs^111^. Additionally, the HEIDI test was performed. HEIDI uses multiple SNPs in a *cis*-mQTL region to distinguish pleiotropy from linkage. As it is described in Zhu et al.2016^45^, under the hypothesis of pleiotropy, where DNAm and a trait share the same causal variant, the *b*_*xy*_ values calculated for any SNPs in LD with the causal variant are identical. Therefore, testing against the null hypothesis that there is a single causal variant is equivalent to testing whether there is heterogeneity in the *b*_*xy*_ values estimated for the SNPs in the *cis*-mQTL region. For each DNAm probe that passed the genome-wide significance (P_nominal_ < 5×10^−8^) threshold for the SMR test, HEIDI tested the heterogeneity in the *b*_*xy*_ values estimated for multiple SNPs in the *cis*-mQTL region. In this analysis, significant pleiotropic associations between DNAm and the neuropsychiatric diseases were selected as those with P_SMR_ corrected Bonferroni < 0.05 and P_HEIDI_ > 0.05 (not showing heterogeneity).

### Colocalization analyses

Recalling the definition of *cis*-mQTLs in our analysis, all DNAm sites with at least one significant mQTL (P_nominal_ < 5×10^−8^) and located within 0.5 Mb of the regions defined in the original neuropsychiatric GWAS were tested. Colocalization analysis was performed as described in Giambartolomei C. et al.2014 with the R *coloc* package^45^. In total 17,341, 5,466, and 4,643 mQTL-CpGs were tested for BIP (n = 63 autosomal regions), MDD (n = 98 autosomal regions) and SCZ (n = 279 autosomal regions), respectively. In both the GWAS data and our mQTLs we imputed the regression coefficients, their variances and the SNP minor allele frequencies, and the prior probabilities were left as their default values. This methodology quantifies the support across the results of each GWAS for five hypotheses by calculating the posterior probabilities, denoted as *PPi* for hypothesis *Hi*.

H_0_: there exist no causal variants for either trait;

H_1_: there exists a causal variant for one trait only, GWAS;

H_2_: there exists a causal variant for one trait only, DNA methylation;

H_3_: there exist two distinct causal variants, one for each trait;

H_4_: there exist a single causal variant common to both traits.

### Conditional analyses

We performed a GCTA conditional analysis^112,113^ conditioning on the top *cis*-mQTL of those probes that passed SMR (P_SMR_ Bonferroni < 0.05) but failed to pass the HEIDI test (P_HEIDI_ < 0.05), due to the fact that heterogeneity may sometimes be driven by real secondary signals. We also performed the conditional analysis using GWAS summary data of the same set of SNPs (SNPs in the *cis*-mQTL region conditioning on the top *cis*-mQTL) for each of the three phenotypes (BP, MDD, SCZ). For any of these regions where there was evidence of a secondary signal (P_conditional_ < 5×10^−8^), in either mQTL or GWAS data, we reran the conditional analyses in both mQTL and GWAS data conditioning on the secondary signal and then used the conditional results for SMR and HEIDI tests. In this step, significant secondary pleiotropic associations between DNAm and the neuropsychiatric diseases were selected as those with Bonferroni-corrected P_SMR_ < 0.05 and P_HEIDI_ > 0.05 (not showing heterogeneity).

### RICHS eQTMs

RICHS recruited mother and infant pairs from March 2009 to May 2013 following delivery at the Women and Infants Hospital of Rhode Island. RICHS selected infants both small for GA, large for GA and controls born appropriate for GA matched on sex, GA (±3 days), and maternal age (±2 years). The study protocol was approved by the Institutional Review Boards of Brown University and Women and Infants Hospital of Rhode Island. Placental RNA-seq data from a subset of samples (n = 200) were obtained using the Illumina Hi-Seq 2500 platform, aligned to the human reference genome and RNA transcript abundance was quantified using Salmon^114^. About 20 million single-end RNA-seq reads were generated on each sample^115^. Placental DNAm data (n = 220) were obtained using the Illumina Infinium HumanMethylation450 BeadChip, preprocessed, and normalized with the minfi R package^116^. eQTMs were calculated by implementing linear models in MatrixEQTL, considering *cis*-windows of Mb up and downstream of each CpG in a total of 195 placenta samples. Linear regressions were adjusted by sex, 5 PCs of expression and the Planet estimated cell types. Results were corrected with FDR.

The gene set enrichment analysis of the eQTM-genes associated to SCZ according to our SMR and colocalization approaches was performed using the Reactome gene list analysis^53^, with the default settings recommended by the developers in the Reactome database release 83 and the pathway browser version 3.7.

### Code availability

The code for the genotype and methylation QC, as well as the TensorQTL nominal mapping is available in this GitHub repository link: https://github.com/ariadnacilleros/Cis-mQTL-mapping-protocol-for-methylome. The rest of the analysis is available in this GitHub repository link:https://github.com/ariadnacilleros/Cilleros-PortetA.etal.

### Data availability

The three complete placental *cis*-mQTL databases are publicly available online in the following address: https://smari.shinyapps.io/shi. To be uploaded to a repository.

## Data Availability

All data produced in the present work are contained in the manuscript or online at:
https://smari.shinyapps.io/shi

## Acknowledgements

We would like to acknowledge all the INMA and RICHS participants and researchers, for their kind collaboration and support.

## Author contributions

N.F.J., J.R.B. and A.C.P conceived and designed the study. Data analyses were carried out by A.C.P., C.L., S.M., C.B. and N.F.J. I.G.S., D.G.M., A.H.L., B.P.G.G. and M.B. helped with the data analysis and provided valuable analytical advice. M.C.T., M.L., A.I., A.B., G.E., R.S.B., L.S.M., J.C., S.L., M.F.F., M.V., J.I., M.G., C.M., M.B. and J.R.B. actively participated in sample recruitment and/or data acquisition. A.C.P., N.F.J. and J.R.B. interpreted the results. N.F.J. and A.C.P. wrote the first draft of the manuscript. N.F.J directed the project. All authors (A.C.P., C.L., S.M., M.C.T., M.L., A.I., A.B., I.G.S., D.G.M., G.E., A.H.L., R.S.B., C.E.B., B.P.G.G., L.S.M., J.C., S. L., M.F.F., M.V., J.I., M.G., C.M., M.B., J.R.B. and N.F.J.) made substantial contributions to the acquisition, analysis, or interpretation of data, and read and critically revised the manuscript.

