## Supplementary Figure File for "Potentially causal associations between placental DNA methylation and schizophrenia and other neuropsychiatric disorders"

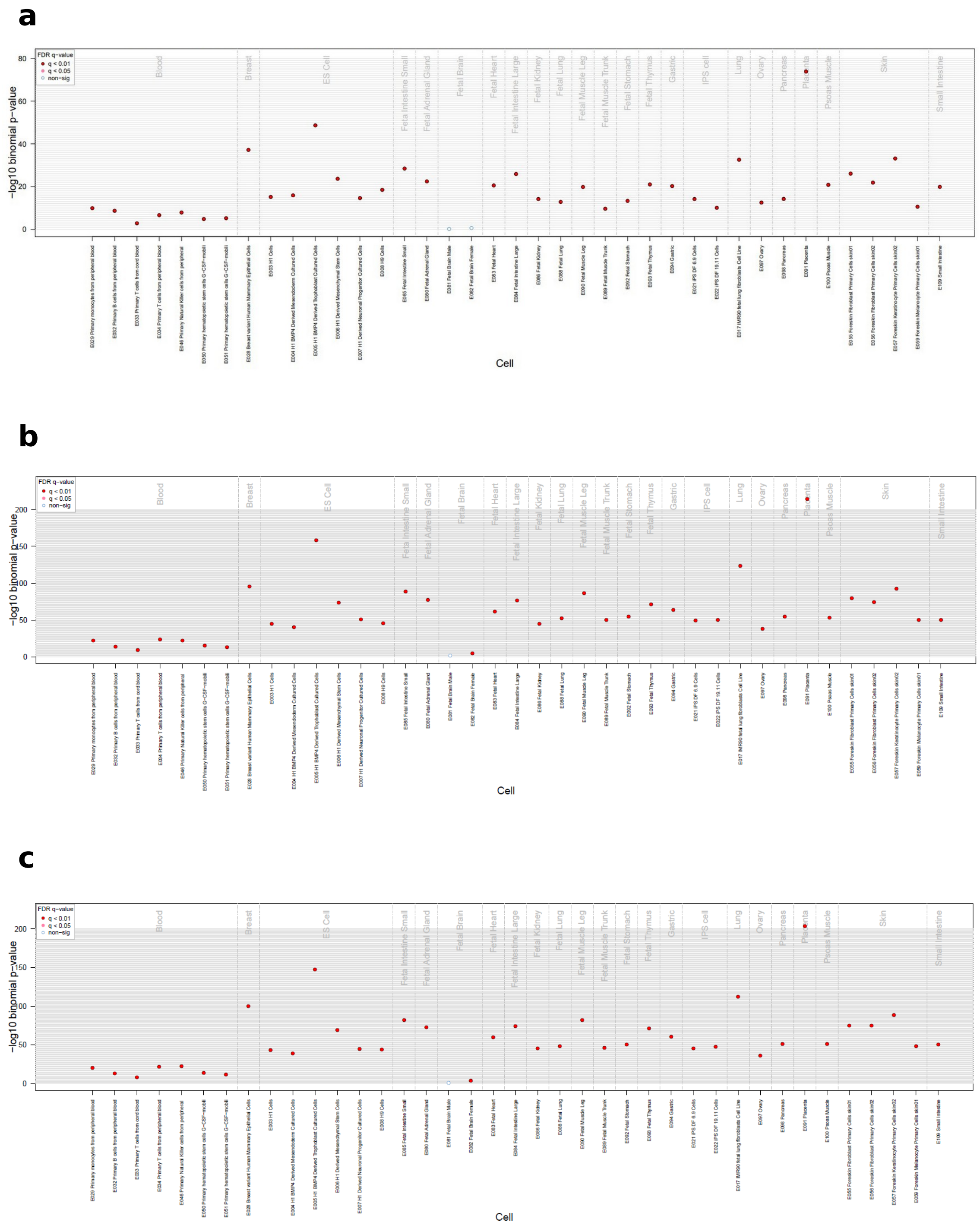


**Supplementary Figure 1. Enrichment on fetal placenta-specific H3K4me1 broadPeaks.** We performed an enrichment analysis of H3K4me1 broadPeaks on the top 10,000 mQTL-CpGs from the **a** nominal, **b** permuted and **c** conditional database with eFORGE. The Y-axis represents the -log10 binomial p-value of the enrichment analysis, while different tissues are shown in the X-axis. False Discovery Rate (FDR) corrected q-values below 0.01 and 0.05 are represented by red and pink dots, respectively, while blue dots show q-values >0.05.

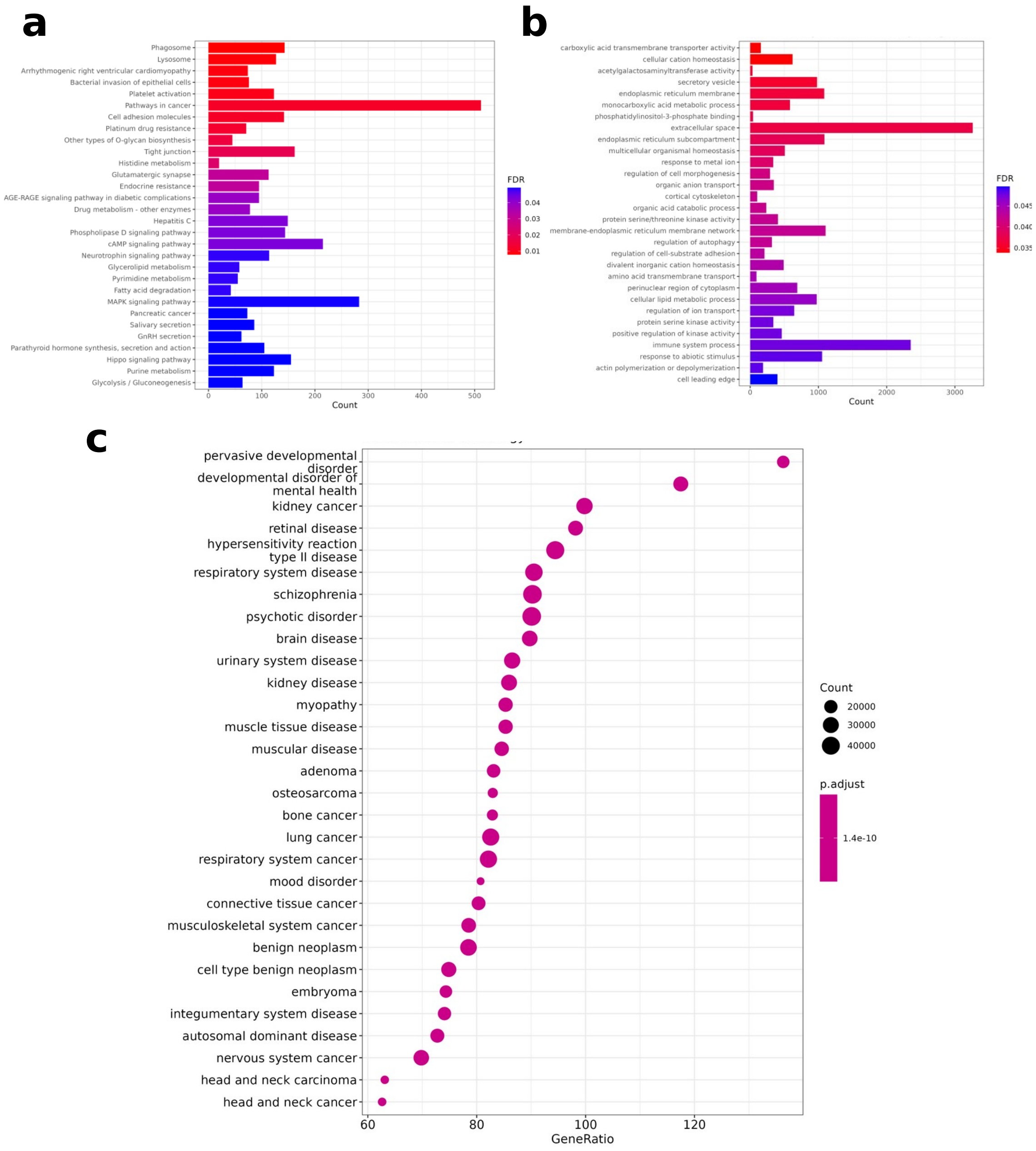


**Supplementary Figure 2. Over-representation and gene set enrichment analysis of the mQTL-CpGs from the nominal database.** An overrepresentation analysis was performed using MissMethyl R package and Gene Ontology (GO) and Kyoto Encyclopaedia of Genes and Genomes (KEGG) gene sets. Results are plotted in the **a** and **b** barplots, respectively. Additionally, a gene set enrichment analysis was performed with the DOSE R package, using the Disease Ontology (DO) database. DOSE results are shown in **c**. In all three plots, the Y-axis represents the top gene sets enriched, and the adjusted p-value is colour coded. In plots **a** and **b**, the X-axes represent the counts, while in **c**, the counts are reported by the dot size and the X-axis represents the gene ratio.

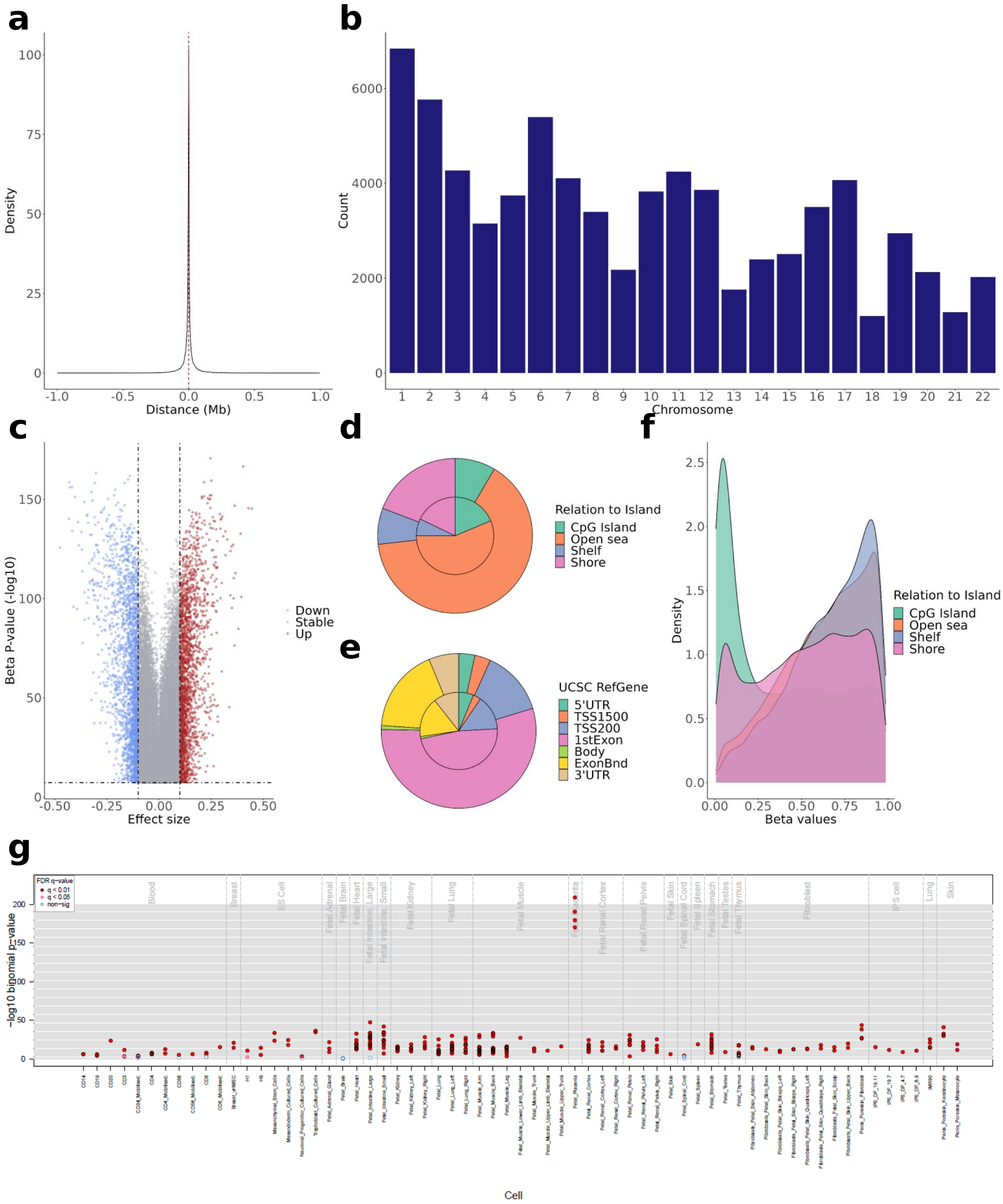


**Supplementary Figure 3. Characterization of the placental *cis*-mQTLs from the permuted database.** The permuted *cis*-mQTL database contains 74,614 mQTLs with 74,614 and 54,294 unique CpGs and SNPs, respectively. The distance between the SNP-CpG pair participating in the *cis*-mQTLs reported is displayed in the **a** density plot, where the X-axis represents the distance in Mb. The red line represents the median SNP-CpG distance of 7,1 kb. The distribution of the *cis*-mQTLs reported along the chromosomes is shown in the **b** barplot, where the X-axis represents the autosomal chromosomes. The uniform distribution of the effect sizes from the *cis*-mQTLs reported is pictured in the **c** volcano plot, where the Y-axis illustrates the -log10 nominal p-value, and the X-axis the effect size. The blue and the red dots show the mQTLs with a negative and a positive effect of the effect allele, respectively. The distribution of the EPIC array probes (inner circle) and the mQTL-CpGs (outer circle) considering the Relation To Island and the UCSC RefGene annotation is displayed in the **d** and **e** piecharts, respectively. The methylation beta values, ranging from 0 to 1, of the mQTL-CpGs stratified by the Relation To Island annotation is shown as the **f** density plot, where methylation values are depicted in the X-axis. The eFORGE enrichment on DNase I hotspots considering the top 10,000 mQTL-CpGs is shown in the **g** plot. The Y-axis represents the -log10 binomial p-value of the enrichment, and the X-axis the tissue. False Discovery Rate (FDR)-corrected q-values below 0.01 and 0.05 are represented by red and pink dots, respectively, while blue dots show q-values >0.05.

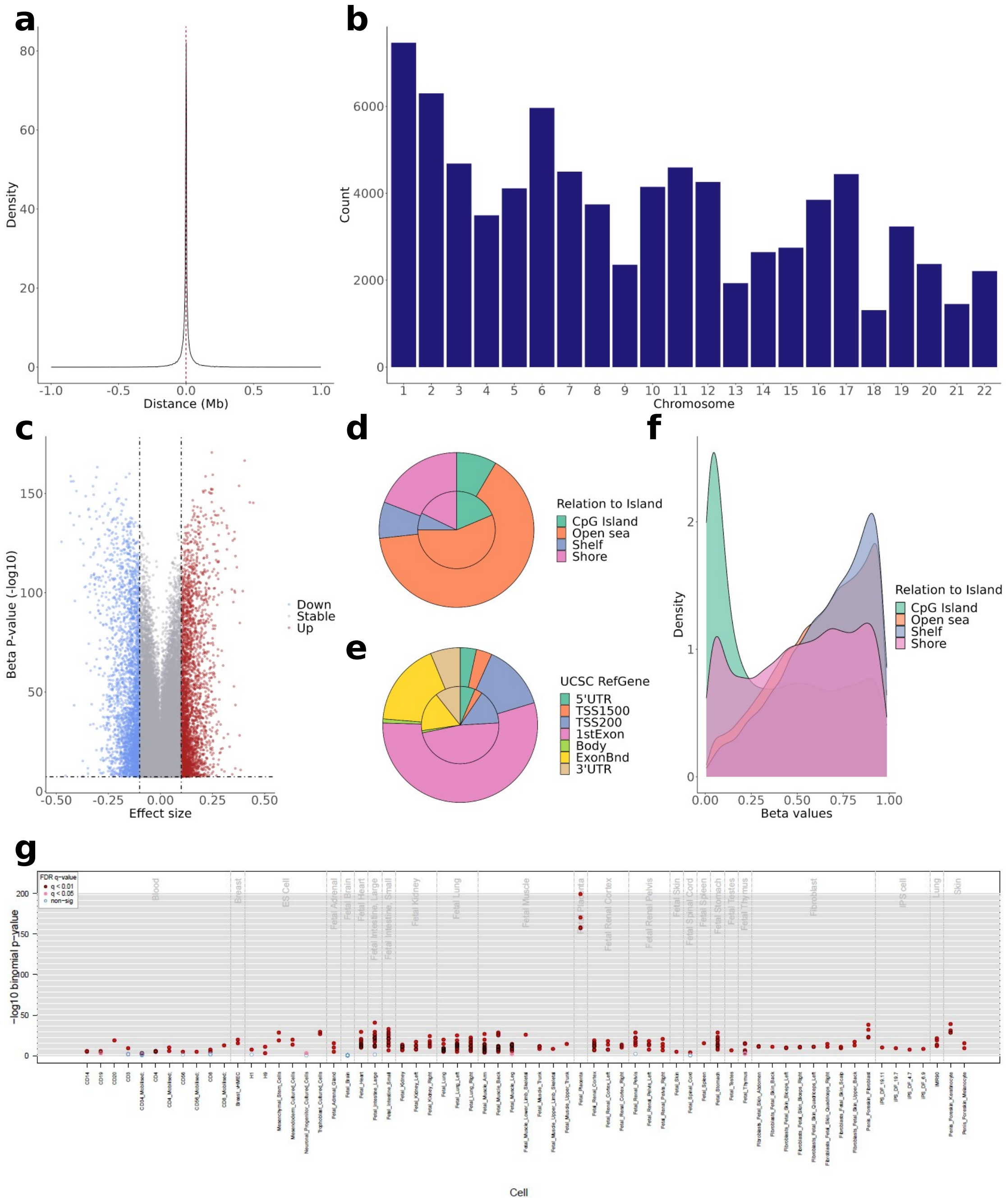


**Supplementary Figure 4. Characterization of the placental *cis*-mQTLs from the conditional database.** The conditional *cis*-mQTL database contains 81,826 mQTLs with 75,241 and 59,957 unique CpGs and SNPs, respectively. The distance between the SNP-CpG pair participating in the *cis*-mQTLs reported is displayed in the **a** density plot, where the X-axis represents the distance in Mb. The red line represents the median SNP-CpG distance of 7,5 kb. The distribution of the *cis*-mQTLs reported along the chromosomes is shown in the **b** barplot, where the X-axis represents the autosomal chromosomes. The uniform distribution of the effect sizes from the *cis*-mQTLs reported is pictured in the **c** volcano plot, where the Y-axis illustrates the -log10 nominal p-value, and the X-axis the effect size. The blue and the red dots show the mQTLs with a negative and a positive effect of the effect allele, respectively. The distribution of the EPIC array probes (inner circle) and the mQTL-CpGs (outer circle) considering the Relation To Island and the UCSC RefGene annotation is displayed in the **d** and **e** piecharts, respectively. The methylation beta values, ranging from 0 to 1, of the mQTL-CpGs stratified by the Relation To Island annotation is shown as the **f** density plot, where methylation values are depicted in the X-axis. The eFORGE enrichment on DNase I hotspots considering the top 10,000 mQTL-CpGs is shown in the **g** plot. The Y-axis represents the -log10 binomial p-value of the enrichment, and the X-axis the tissue. False Discovery Rate (FDR) corrected q-values below 0.01 and 0.05 are represented by red and pink dots, respectively, while blue dots show q-values >0.05.

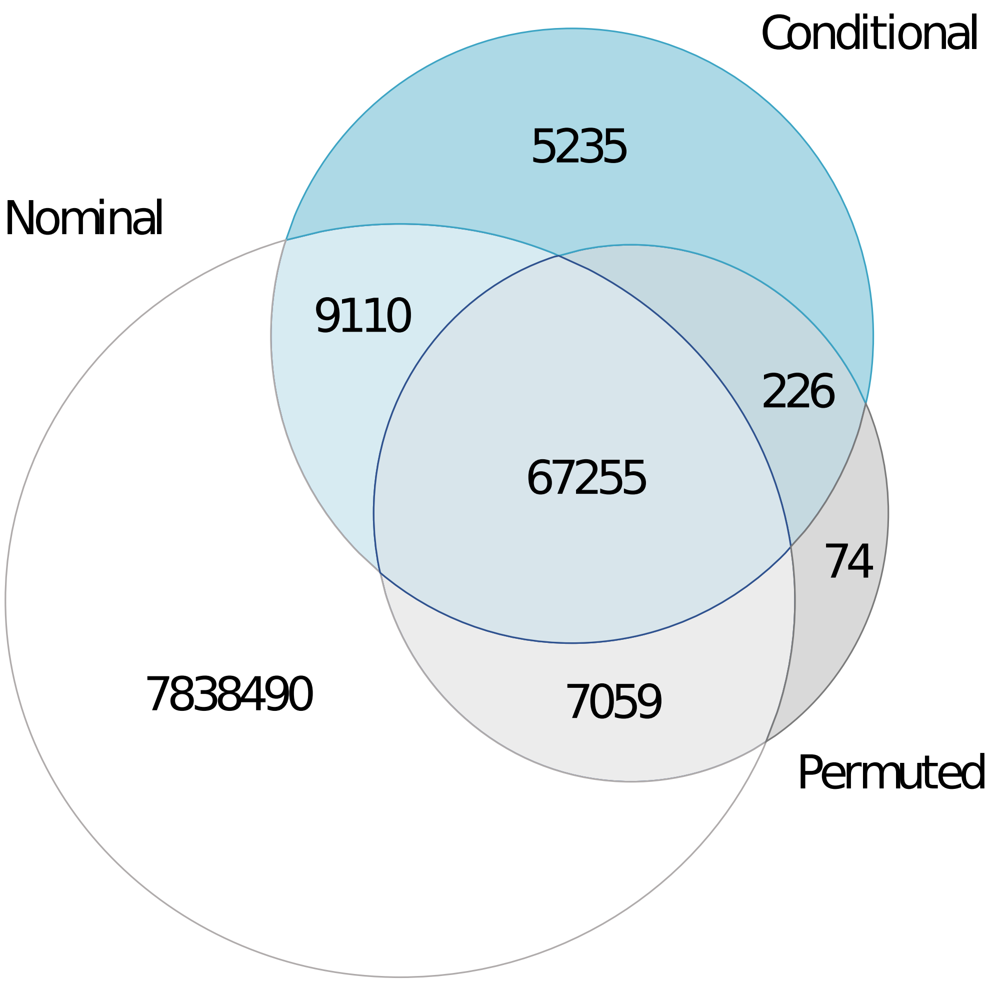


**Supplementary Figure 5. Overlap between the nominal, conditional and permuted databases.** The overlap between the mQTLs (CpG-SNP pairs) is shown in a Venn diagram.

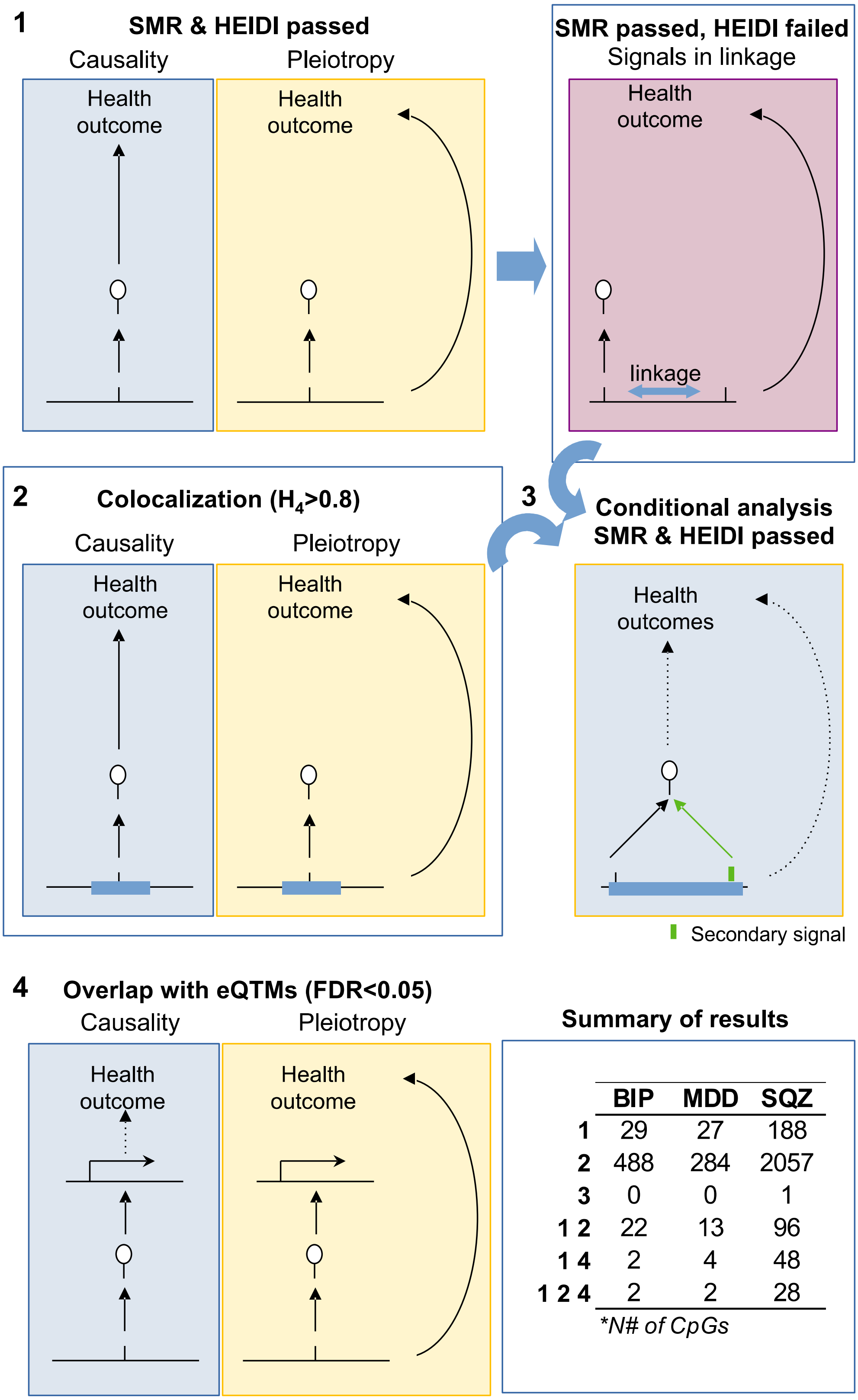


| | BIP | MDD | SQZ |
|---|---|---|---|
| 1 | 29 | 27 | 188 |
| 2 | 488 | 284 | 2057 |
| 3 | 0 | 0 | 1 |
| 1 2 | 22 | 13 | 96 |
| 1 4 | 2 | 4 | 48 |
| 1 2 4 | 2 | 2 | 28 |

**N# of CpGs*

**Supplementary Figure 6. Workflow and results of the main analyses performed.** In a first step, we performed the SMR and HEIDI tests, followed by colocalization. Those hits that passed SMR and colocalization, but failed to pass HEIDI test, were considered for a conditional analysis for a second round of SMR and HEIDI tests, with the aim of discovering secondary, independently associated signals. Finally, those CpGs passing the SMR and HEIDI tests, were interrogated in the eQTMs database of the RICHS cohort. The CpG overlap between the four approaches is summarized in the right-bottom table.

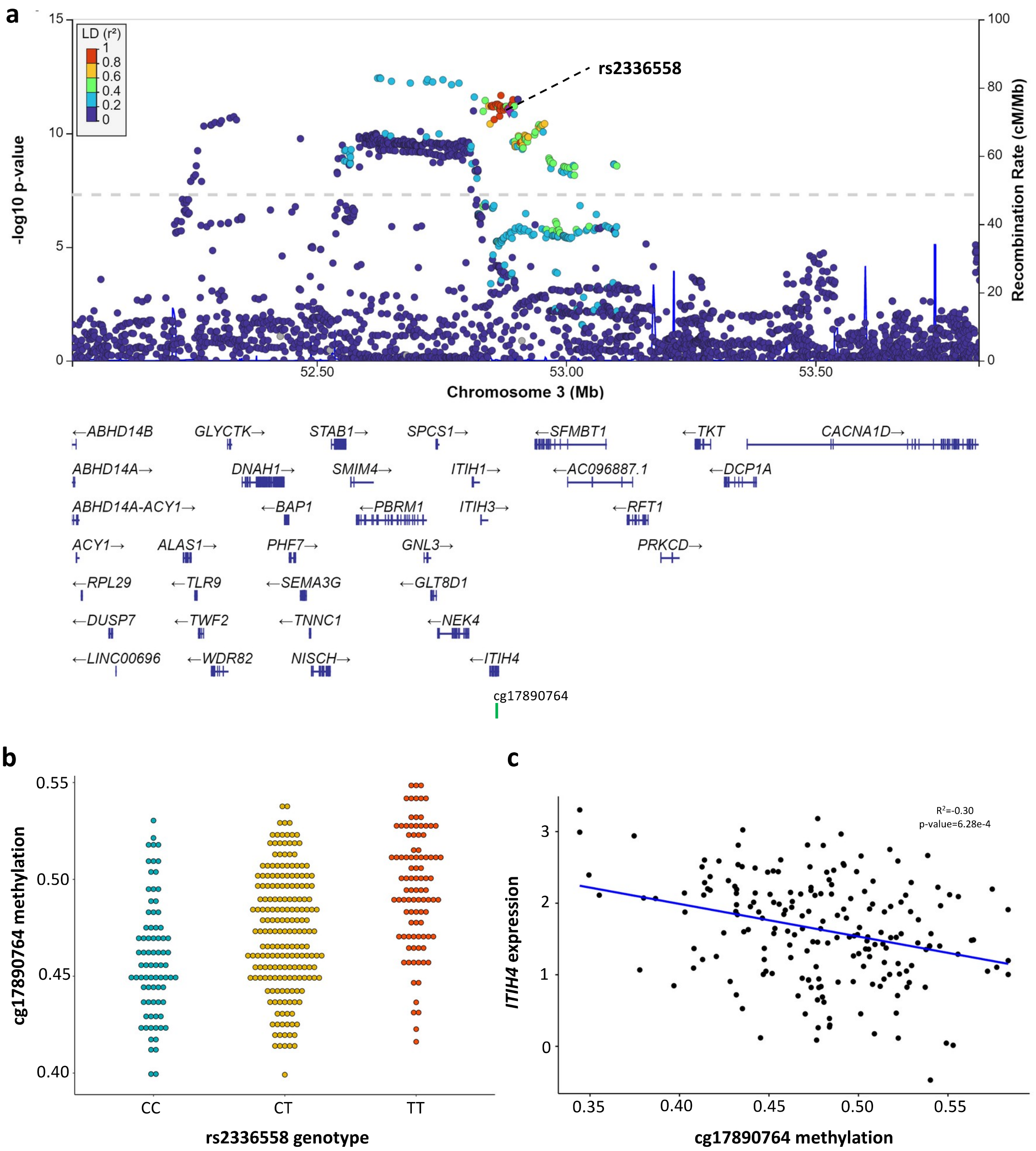


**Supplementary Figure 7. cg17890764, the mQTL-CpG associated with BIP according to SMR is a placental eQTM of *ITIH4*.** The genomic region of the rs2336558 mQTL-SNP, highlighted as a purple diamond, in the original BIP GWAS is represented in the **a** locusZoom plot. The X-axis displays the genomic region of chromosome 3 in Mb, the distribution of the coding genes, and the mQTL-CpG, cg17890764. The Y-axis represents the -log10 p-value from the original GWAS, and the SNPs are colour coded as a function of the linkage disequilibrium with the highlighted SNP. The significant mQTL, rs2336558-cg17990764, is plotted in the **b** dotplot, where the X-axis represents the beta methylation values of cg17990764, ranging from 0 to 1. The Y-axis displays the genotype of rs2336558. The cg17990764-*ITIH4* eQTM is shown in **c**, where the X-axis represents the methylation values of the CpG, ranging from 0 to 1, and the Y-axis displays the placental expression of the *ITIH4* gene.

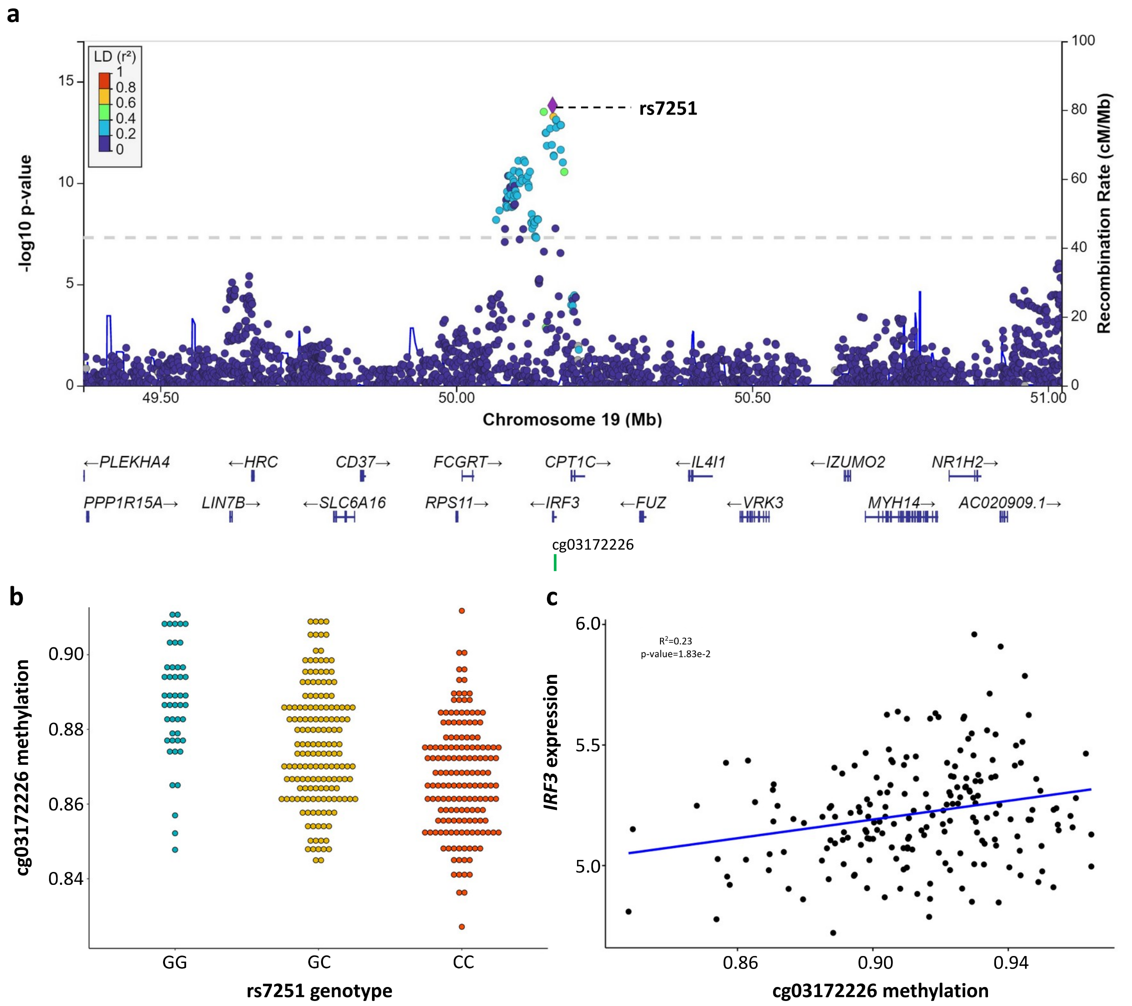


**Supplementary Figure 8. cg03172226, the mQTL-CpG associated with SCZ according to SMR is a placental eQTM of *IRF3*.** The genomic region of the rs7251 mQTL-SNP, highlighted as purple diamond, in the original SCZ GWAS is represented in the **a** locusZoom plot. The X-axis displays the genomic region of chromosome 19 in Mb, the distribution of the coding genes, and the mQTL-CpG, cg03172226. The Y-axis represents the -log10 p-value from the original GWAS, and the SNPs are colour coded as a function of the linkage disequilibrium with the highlighted SNP. The significant mQTL, rs7251-cg03172226, is plotted in the **b** dotplot, where the X-axis represents the beta methylation values of cg03172226, ranging from 0 to 1. The Y-axis displays the genotype of rs7251 SNP. The cg03172226-*IRF3* eQTM is shown in **c**, where the X-axis represents the methylation values of the CpG, ranging from 0 to 1, and the Y-axis displays the placental expression of the *IRF3* gene.

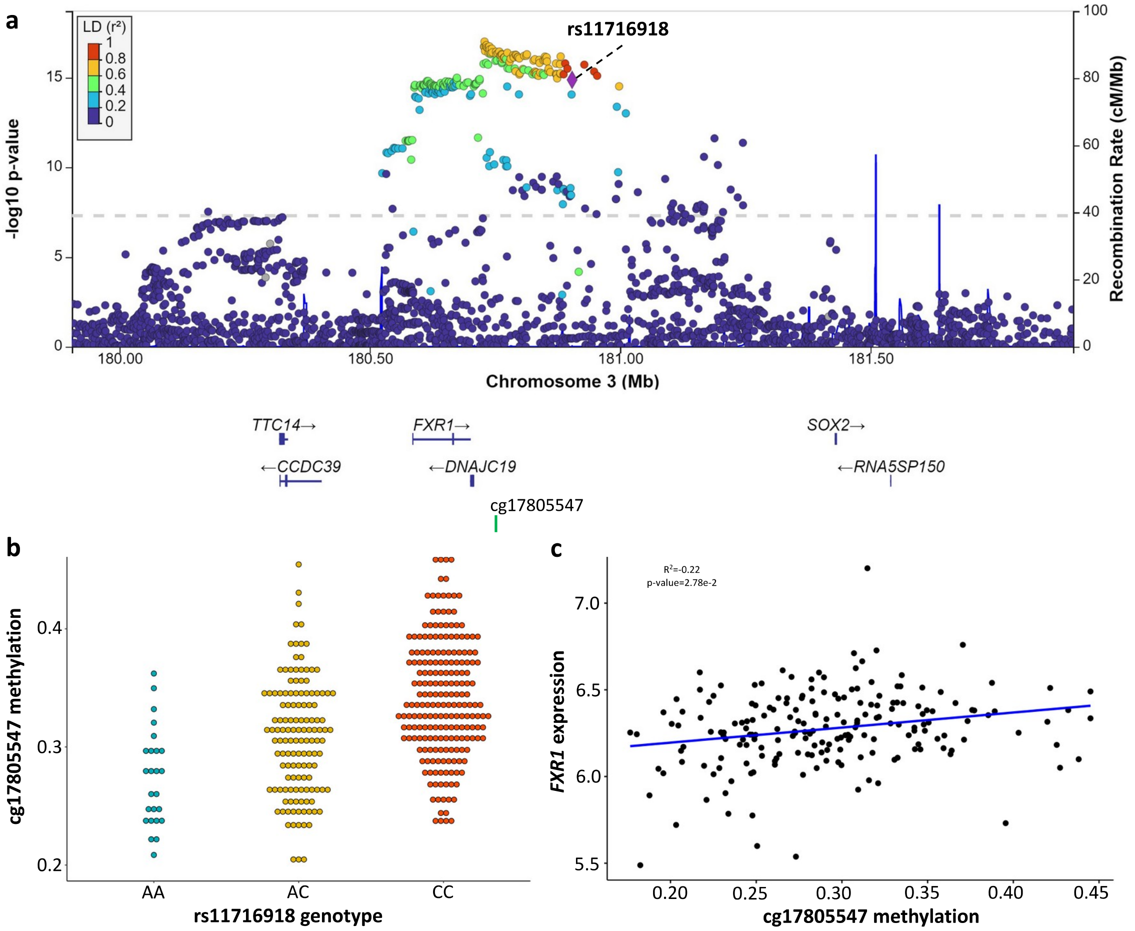


**Supplementary Figure 9. cg17805547, the mQTL-CpG associated with SCZ according to SMR is a placental eQTM of *FXR1*.** The genomic region of the rs11716918 mQTL-SNP, highlighted as a purple diamond, in the original SCZ GWAS are represented in the **a** locusZoom plot. The X-axis displays the genomic region of chromosome 3 in Mb, the distribution of the coding genes, and the mQTL-CpG, cg17805547. The Y-axis represents the -log10 p-value from the original GWAS, and the SNPs are colour coded as a function of the linkage disequilibrium with the highlighted SNP. The significant mQTL, rs11716918-cg17805547, is plotted in the **b** dotplot, where the X-axis represents the beta methylation values of cg17805547, ranging from 0 to 1. The Y-axis displays the genotype of rs11716918 SNP. The cg17805547-*FXR1* eQTM is shown in **c**, where the X-axis represents the methylation values of the CpG, ranging from 0 to 1, and the Y-axis displays the placental expression of the *FXR1* gene.

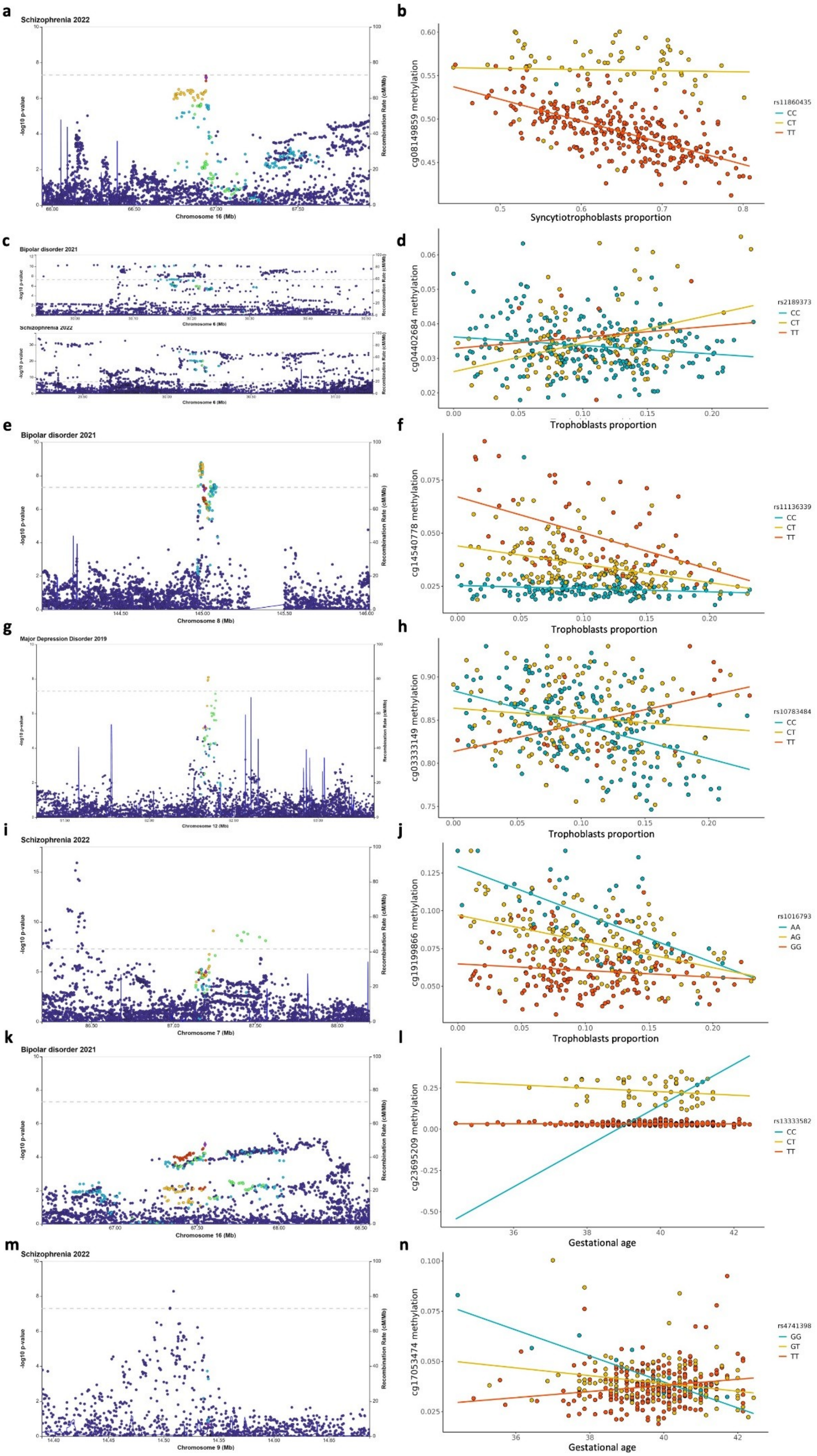
a
Schizophrenia 2022
-log10 p-value
Recombination Rate (cM/Mb)
Chromosome 16 (Mb)
b
cg08149859 methylation
Syncytiotrophoblasts proportion
rs11860435
CC
CT
TT
c
Bipolar disorder 2021
Chromosome 6 (Mb)
Schizophrenia 2022
Chromosome 6 (Mb)
d
cg04402684 methylation
Trophoblasts proportion
rs2189373
CC
CT
TT
e
Bipolar disorder 2021
Chromosome 8 (Mb)
f
cg14540778 methylation
Trophoblasts proportion
rs11136339
CC
CT
TT
g
Major Depression Disorder 2019
Chromosome 12 (Mb)
h
cg03333149 methylation
Trophoblasts proportion
rs10783484
CC
CT
TT
i
Schizophrenia 2022
Chromosome 7 (Mb)
j
cg19199866 methylation
Trophoblasts proportion
rs1016793
AA
AG
GG
k
Bipolar disorder 2021
Chromosome 16 (Mb)
l
cg23695209 methylation
Gestational age
rs13333582
CC
CT
TT
m
Schizophrenia 2022
Chromosome 9 (Mb)
n
cg17053474 methylation
Gestational age
rs4741398
GG
GT
TT

**Supplementary Figure 10. Significant SMR hits with the imQTLs in BIP, MDD and SCZ.** Significant SCZ SMR hit from STB-imQTLs, rs11860435-cg08149859, is plotted in **a** and **b** locusZoom and dotplot, respectively. Significant BIP, MDD and SCZ SMR hits from TB-imQTLs, rs2189373-cg04402684, rs11136339-cg14540778, rs10783484-cg03333149, and rs1016793-cg19199866, are plotted in **c**, **d**, **e**, **f**, **g**, **h**, **i** and **j** locusZooms and dotplots, respectively. Significant BIP and SCZ SMR hits from GA imQTLs, rs13333582-cg23695209 and rs4741398-cg17053474, are plotted in **k**, **l**, **m** and **n** locusZooms and dotplots, respectively. In all locusZooms, the X-axes display the genomic region involved in Mb, and the Y-axes represent the -log10 p-values from the original GWAS used. Each imQTL-SNP is highlighted as a purple diamond, and the surrounding SNPs are colour coded as a function of the linkage disequilibrium with the highlighted SNP. In all dotplots, the X-axes represent the beta methylation values of each imQTL-CpG, ranging from 0 to 1. And the Y-axes display the genotype of each imQTL-SNP.
